# Role of Statins in the Primary Prevention of Atherosclerotic Cardiovascular Disease and Mortality in the Population with Mean Cholesterol in the Near-Optimal to Borderline High Range: A Systematic Review and Meta-Analysis

**DOI:** 10.1101/2020.10.02.20205849

**Authors:** Bishnu M. Singh, Hari K. Lamichhane, Sanjay S. Srivatsa, Prabhat Adhikari, Bikash J. Kshetri, Sijan Khatiwada, Dhan B. Shrestha

## Abstract

**Objective:** The objective of this meta-analysis was to analyze the benefits and harms of treating the population with statins in those having mean low-density lipoprotein cholesterol (LDL-C) in the near-optimal (100 to 129 mg/dl) to borderline high (130 to 159 mg/dl) range and free of cardiovascular disease (CVD). Methods: We searched PubMed, PubMed Central, Cochrane Library, and Google Scholar databases for randomized controlled trials (RCTs) published between 1994 and July 2020. We included RCTs with greater than 90% of participants free of CVD. Two reviewers independently screened the articles using the Covidence software, assessed the methodological quality using the risk of bias 2 tool, and analyzed the data using the RevMan 5.4 software. Results: Eleven trials were included. Statin therapy was associated with a decreased risk of myocardial infarction (RR=0.56, 95% CI: 0.47 to 0.67), major cerebrovascular events (RR=0.78, 95% CI: 0.63 to 0.96), major coronary events (RR=0.67, 95% CI: 0.57 to 0.80), composite cardiovascular outcome (RR=0.71, 95% CI: 0.62 to 0.82), revascularizations (RR=0.65, 95% CI: 0.57 to 0.74), angina (RR=0.76, 95% CI: 0.63 to 0.92) and hospitalization for cardiovascular causes (RR=0.74, 95% CI: 0.64 to 0.86). There was no benefit associated with statin therapy for cardiovascular mortality and coronary heart disease mortality. All-cause mortality benefit with statin therapy was seen in the population with diabetes and increased risk of CVD. Statin therapy was associated with no significant increased risk of myalgia, creatine kinase elevation, rhabdomyolysis, myopathy, incidence of any cancer, incidence of diabetes, withdrawal of the drug due to adverse events, serious adverse events, fatal cancer, and liver enzyme abnormalities. Conclusion: Statin therapy was associated with a reduced risk of cardiovascular disease and procedures without increased risk of harm in populations with mean LDL-C near-optimal to the borderline high range without prior atherosclerotic cardiovascular disease.

## 1. Introduction

Atherosclerotic Cardiovascular disease (ASCVD) encompasses four major diseases, including coronary heart disease (CHD), cerebrovascular disease, peripheral artery disease (PAD), and aortic atherosclerosis. It is a major cause of death around the world, contributing about 30% of the total figure [1]. Every year, 17 million deaths, with 7.6 million due to myocardial infarction (MI) and 5.7 million due to stroke, cost the world an amount of 863 billion USD [1]. In the US, this disease costs around 1-3 % of the GDP, and the American Heart Association (AHA) in 2010 estimated the annual cost of cardiovascular disease (CVD) in the US to be around 503.2 billion USD [2]. In the current scenario, CVD has peaked as a growing burden, even in low and middle-income countries that traditionally emphasized undernutrition and infectious diseases.

CVD has different risk factors. Of these, the average cholesterol level of a particular population is a very important factor that determines the ASCVD risk of that population [1]. Previous prospective observational studies revealed that the relationship between serum cholesterol and CHD is a continuously graded one rather than just a threshold one and the risk gradient to be continuous over the whole range of cholesterol concentrations [3]. A study done in Shanghai, China suggested that cholesterol is still an important cause of CHD, where the mean baseline serum cholesterol concentration is considered a normal or low concentration by Western Standards [4]. A study conducted in India revealed that low serum cholesterol has a strong positive relationship with coronary artery disease. It did not show any evidence of a threshold. It concluded that there might be a benefit if serum cholesterol is decreased below the range of what is considered desired in developed nations [5]. Hence, it is essential to study the effect of cholesterol-lowering on cardiovascular prevention in populations with low mean cholesterol.

Among the different types of cholesterol, low-density lipoprotein cholesterol (LDL-C) plays a crucial role in the development of ASCVD. LDL-C is responsible for the development and progression of atheroma and plaque, which upon rupture results in catastrophic cardiovascular events (CVEs) [1]. Statins are cholesterol-lowering drugs that reduce LDL-C concentration by decreasing its synthesis in the liver and increasing its removal from the circulation [6]. There is clear cut evidence of statins in the prevention of cardiovascular events (CVEs) or mortality in those who had prior CVD and those with high cholesterol but without prior CVD. In these populations, the benefits could be well explained by the cholesterol-lowering effect of statins as well as a group of “cholesterol-independent” or “pleiotropic “effects, which includes improvement in the functioning of endothelial cells, increased stability of atherosclerotic plaques, reduced oxidative stress and inflammation, inhibition of vascular smooth muscle proliferation and platelet aggregation [7]. However, there is limited evidence of the role of statins in the prevention of cardiovascular disease in the population with average cholesterol in the near-optimal (LDL-C: 100 to 129 mg/dl) to borderline high (LDL-C: 130 to 159 mg/dl) range.

The main objective of this meta-analysis was to analyze the benefits and harms of treating the population without CVD, with mean LDL-C in the near-optimal to borderline high range, and provide clear evidence about its role in this population to the scientific community. This meta-analysis, based on primary preventive randomized controlled trials (RCTs) of statins, will be helpful for guideline makers and clinicians to know the effects of treating the population with statins for primary prevention of cardiovascular disease, even though the population has mean LDL-C in the near-optimal to borderline high range.

## 2. Methods

We followed the guidelines of the Preferred Reporting Items for Systematic reviews and Meta-Analyses (PRISMA 2009) for conducting the meta-analysis [8].

### 2.1 Study Protocol

We did preliminary searches and literature reviews on our research question. We then prepared our protocol according to the guidelines of the Preferred Reporting Items for Systematic review and Meta-Analysis Protocols 2015 (PRISMA-P 2015). We published our protocol at the Research Registry on July 13, 2020, with a unique identifying number: reviewregistry946.

### 2.2 Search Strategy

We used electronic databases like PubMed, Cochrane Library, PubMed Central (PMC), and Google Scholar for searching relevant articles to answer our research question from January 1, 1994, to July 2020. We customized our search to include any clinical trial or review articles that had stated the role of statins in the primary prevention of cardiovascular events or mortality. We searched for English language studies conducted in human subjects. The search strategies for different electronic databases are shown in Supplementary Material 1.

### 2.3 Study Selection

After we completed our search, we imported all articles in the Mendeley software. We removed the duplicates in the Mendeley software and exported the file. This file was then imported into the Covidence software. The Covidence software removed duplicates as well. Two reviewers (B.M.S. and H.K.L.) independently screened the articles based on their titles and abstracts. The same reviewers again did full-text screening independently. Titles, abstracts, and full-text screening were done using the Covidence software. All conflicts were resolved by other authors (P.A. and S.S.S.). At every step, we used our eligibility criteria to screen and finally selected the studies included in our meta-analysis. The inclusion criteria were: (i) randomized clinical trial comparing statin with placebo, standard therapy or no treatment; (ii) follow-up of at least one year; (iii) > 90% of participants free of CVD to ensure the treatment effect on the primary prevention population or studies reporting data separately in the subgroup who did not have CVD and provide specific numbers for participants and events in that subgroup; (iv) average LDL-C of participants between 100 to 159 mg/dl; (v) 100 participants in the intervention group; and (vi) studies reporting one or more of the following outcomes: cardiovascular events (CVEs), coronary heart disease (CHD) events or death, all-cause mortality, unstable angina, acute myocardial infarction (fatal or non-fatal), stroke or transient ischemic attack (fatal or non-fatal), surgical or percutaneous revascularization, and heart failure. The exclusion criteria were:(i) studies not involving RCT; (ii) studies that investigated and reported only statin-related nonclinical and intermediate surrogate endpoints such as carotid intima media thickness changes, lipid levels, or angiographic outcomes; (iii) studies done on specific groups like late chronic kidney disease (CKD), renal transplant, hemodialysis, human immunodeficiency virus (HIV) or aortic stenosis patients; (iv) studies done on diabetic patients with glycated hemoglobin (HbA1C) > 12%, participants whose predicted 10-year risk of a major coronary event or stroke exceeded approximately 20%, studies prescreened participants for atherosclerosis using ultrasound; (v) lack of a statin-free control group in the study design, compared high to low dose statins; and (vi) studies not reporting the proportion of participants free of CVD. Because of our LDL-C criteria, we also excluded studies done in subjects with familial hyperlipidemia.

### 2.4 Data Extraction

We extracted the following data from the studies included in our meta-analysis: (i) year of publication; (ii) participants in statins and control groups; (iii) duration of follow-up; (iv) percentage of participants with prior CVD; (v) participant characteristics like mean age, percentage of diabetes, percentage of the current smoker, percentage of women, mean BMI, race, mean SBP/DBP; (vi) type and the dose of statin; (vii) mean baseline level of total cholesterol (TC), LDL-C; (viii) the percentage of participants with a family history of premature CHD; (ix) target population; (x) study design; (xi) method and mode of statistical analysis; (xii) endpoints of study; and (xiii) adverse events noted among participants. We also extracted the data for the different predetermined outcome measures. Three reviewers (B.M.S., H.K.L., and D.B.S.) extracted the data independently, and any conflicts during the process of data extraction were resolved by other authors (P.A. and S.S.S.).

### 2.5 Outcome Measures

Primary outcomes were myocardial infarction (MI), major cerebrovascular and coronary events, all-cause mortality, composite cardiovascular outcomes, CHD mortality, cardiovascular mortality, muscle-related adverse events, the incidence of any cancer, and incidence of diabetes. Secondary outcomes were revascularizations, angina, hospitalizations for cardiovascular causes, withdrawal from the drug due to adverse events, serious adverse events, fatal cancer, and liver enzyme abnormalities.

### 2.6 Quality Assessment

We assessed the quality of the included studies by using the Cochrane quality assessment tool for RCTs [9]. We analyzed the risk of bias in each RCT using the risk of bias 2 (RoB 2) tool under the following headings: random sequence generation (selection bias), allocation sequence concealment (selection bias), blinding of participants and personnel (performance bias), blinding of outcome assessment (detection bias), incomplete outcome data (attrition bias), selective outcome reporting (reporting bias) and other potential sources of bias. Depending on the risk of bias, the tool rated RCTs as “Low risk,” “Unclear risk,” and “High risk.” Two reviewers (B.M.S. and H.K.L.) independently assessed the risk of bias using the RoB 2 tool. Any disagreements were resolved by other authors (P.A. and S.S.S.).

### 2.7 Statistical Analyses

We calculated the risk ratios (RR), odds ratio (OR), and 95% confidence interval (CI) of the events occurring in the statin and placebo groups according to predefined outcomes in our protocol. Statistical analysis was done using the RevMan 5.4 software. We used the Mantel-Haenszel statistical method to measure the effect size. We used the fixed/random-effects model for the pooling of studies as per the heterogeneity. We assessed the heterogeneity using the I-squared (I^2^) test and used the Cochrane Handbook for Systematic Reviews of Interventions for interpretation of I-squared (I^2^) test as follows: “0% to 40%: might not be important; 30% to 60% may represent moderate heterogeneity; 50% to 90%: may represent substantial heterogeneity; 75% to 100%: considerable heterogeneity.” When significant heterogeneity was present, we explored the reasons by analyzing the clinical characteristics of participants, study design, and interventions of the included RCTs. We also tried to explain the heterogeneity in the outcomes by performing sensitivity analysis wherever possible. We did an additional sensitivity analysis to determine the influence on the effect size of the outcomes based on the following criteria: (i) repeating the analysis separately for the population with mean LDL-C in the near-optimal and borderline high range; (ii) repeating the analysis separately for trials with participants > 3000 and < 3000; (iii) repeating the analysis separately for trials with adequate and inadequate randomization; (iv) repeating the analysis separately for trials with blinded and unblinded participants; (v) repeating the analysis separately for trials with average follow-up > 3.5 years and < 3.5 years. We checked for publication bias by drawing a funnel plot for the outcomes that included ten or more RCTs. We visually examined any asymmetry of the funnel plot.

### 2.8 Strength of the Body of Evidence

We assessed the quality of evidence for important outcomes using the Grading of Recommendations Assessment, Development, and Evaluation (GRADE) approach. We evaluated the quality of evidence across the domains of risk of bias, consistency, directness, precision, and publication bias. We assessed the strength of evidence of important outcomes using GRADEpro GDT software. Based on the results, we graded our confidence in the estimate of effect size for outcomes as high (further research unlikely to change our confidence), moderate (further research can have an important impact), low (further research is very likely to have an important impact) or very low (very uncertain in the estimate of effect).

## 3. Results

### 3.1 Search Results

Our search strategy initially identified 16575 articles (PMC 5362, PubMed 4911, Cochrane Library 5002, Google Scholar 1297, and 3 articles from the clinical conference). We imported all articles to the Mendeley software. We removed 450 duplicate articles and 1126 irrelevant articles using the Mendeley software. We imported 14999 articles into the Covidence software. Covidence software removed 235 duplicates. We screened 14764 articles by title and abstract and removed 13848 irrelevant articles. We selected 906 articles for full-text screening. We used the Covidence software for title, abstract, and full-text screening. The values of proportionate agreement for the title, abstract, and full-text screening were 0.95 and 0.99, respectively. We obtained the full text of 916 articles and checked their eligibility based on the inclusion and exclusion criteria in the protocol. We excluded 905 articles that could not fit in our eligibility criteria, and finally, 11 RCTs were included for both qualitative and quantitative analyzes [10-20]. Figure 1 shows the PRISMA flow diagram of the study search process.

**Figure 1:**
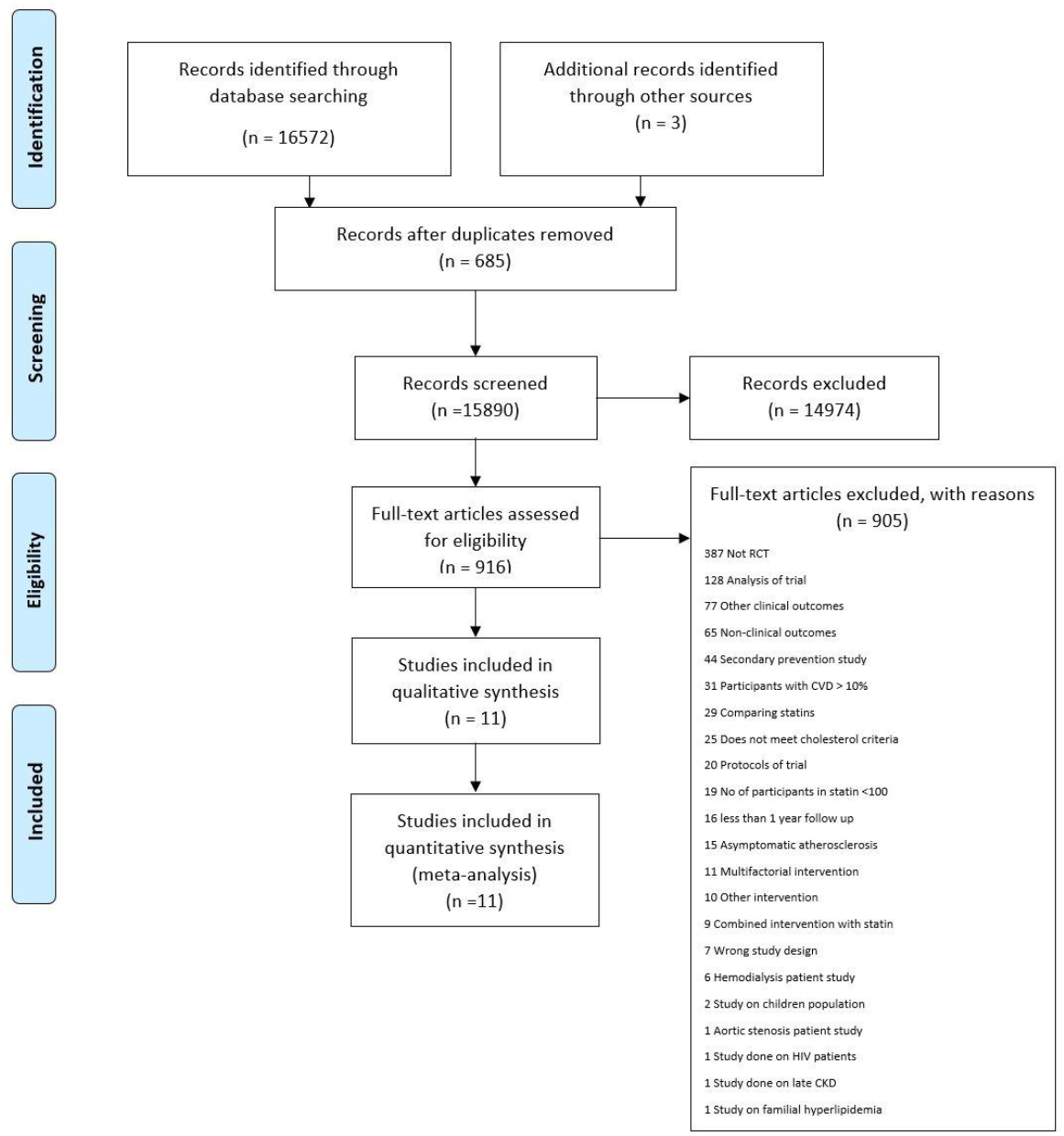
PRISMA flow diagram of the study search process. RCT indicates randomized controlled trial; CVD, cardiovascular disease; HIV, human immunodeficiency virus; CKD, chronic kidney disease.

### 3.2 Characteristics of Included Trials

Eleven RCTs with 58504 participants (29235 in the statin group and 29269 controls) were included: HYRIM [10] (Hypertension High-Risk Management trial), PREVEND IT [11](Prevention of Renal and Vascular Endstage Disease Intervention Trial), Beishuizen et al. [12], CARDS [13] (Collaborative Atorvastatin Diabetes Study), AFCAPS/TexCAPS [14] (Air Force/Texas Coronary Atherosclerosis Prevention Study), ALLHAT-LLT [15] (Antihypertensive and Lipid-Lowering Treatment to Prevent Heart Attack), TRACE RA [16] (Trial of Atorvastatin for the primary prevention of Cardiovascular Events in Rheumatoid Arthritis), MEGA [17] (Management of Elevated Cholesterol in the Primary Prevention Group of Adult Japanese), JUPITER [18] (Justification for the Use of Statins in Prevention: An Intervention Trial Evaluating Rosuvastatin), PROSPER [19] (PROspective Study of Pravastatin in the Elderly at Risk), and HOPE-3 trial [20] (Heart Outcomes Prevention Evaluation). The shortest follow-up was in JUPITER^18^ with a median of 1.9 years, and the longest follow-up was in the HOPE-3 trial [20] with a median of 5.6 years. Supplementary Table 1 shows the information regarding the baseline demographic characteristics of the participants, along with the interventions used in the included trials. Supplementary Table 2 presents the study design, and the target population of the different trials included in this quantitative analysis.

### 3.3 Risk of Bias Assessment

All eleven trials were fully or partially supported by different pharmaceutical companies. Two trials, ALLHAT-LLT [15] and MEGA [17], did not blind the participants and study personnel. Two trials, AFCAPS/TexCAPS [14] and HYRIM [10], did not clearly report the randomization sequence and allocation concealment. In Beishuizen et al., there was a significant loss to follow-up [12]. All adverse events were not reported by the MEGA trial [17]. Figure 2 shows the quality of the included trials assessed by the Risk of Bias 2 (RoB 2) tool [9].

**Figure 2:**
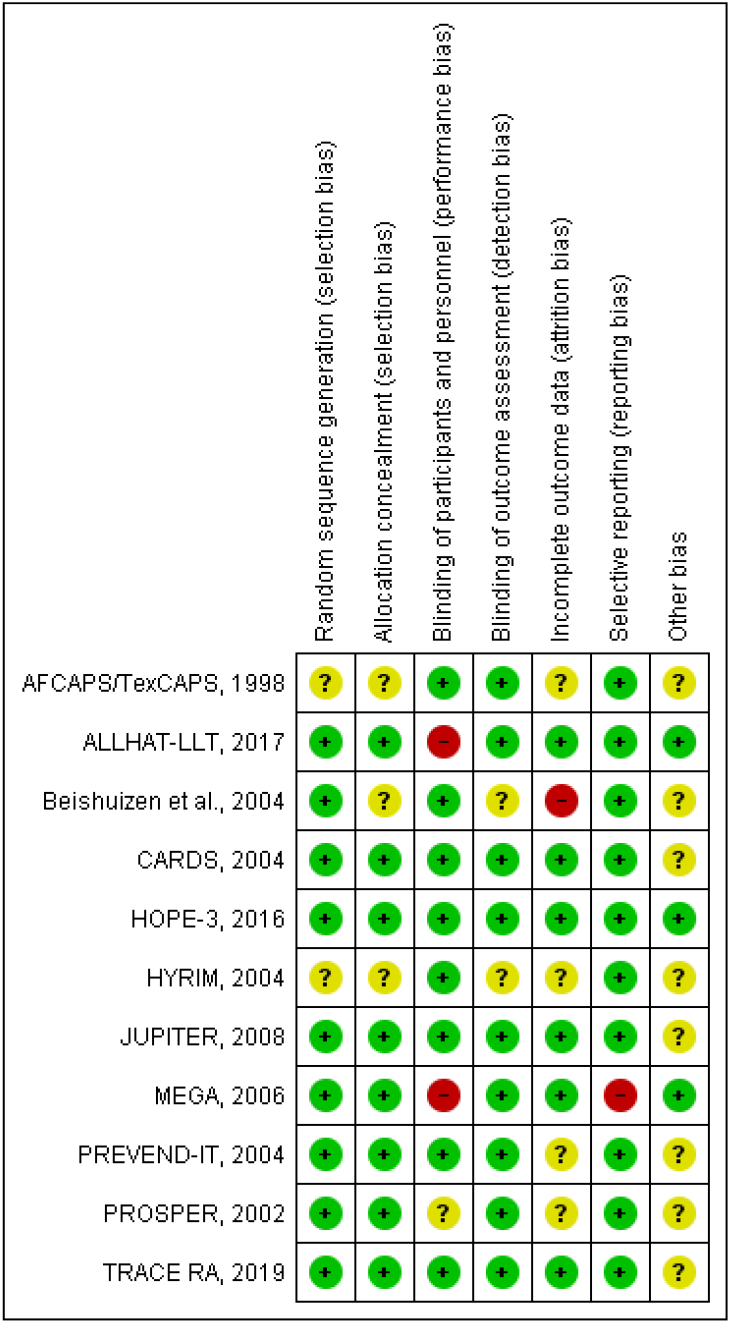
Risk of bias summary. Review authors’ judgments about each risk of bias item for each included study.

### 3.4 Primary Outcomes

#### (1) Myocardial Infarction

This is the composite outcome that included any MI, fatal MI, or nonfatal MI. Six trials [13,14,16-18,20] with 50784 participants, representing 86.8% of the total population, reported MI. AFCAPS/TexCAPS [14] reported fatal and nonfatal MI, CARDS [13] reported occurrence of the first event as fatal or nonfatal MI, HOPE-3 [20] reported MI, JUPITER [18] reported any MI, MEGA [17] reported MI, and TRACE RA [16] reported only non-fatal MI. CARDS [13] reported fatal MI and nonfatal MI separately based on the occurrence of the first event. During the follow-up period, 194/25364 (0.76%) developed MI in the statin group compared with 346/25420 (1.36%) in the control group. Remarkably, participants in the statin group had a lower occurrence of MI in comparison with those of the control group (RR = 0.56, 95% CI: 0.47 to 0.67, I^2^ = 0%), as can be seen in Figure 3.

**Figure 3:**
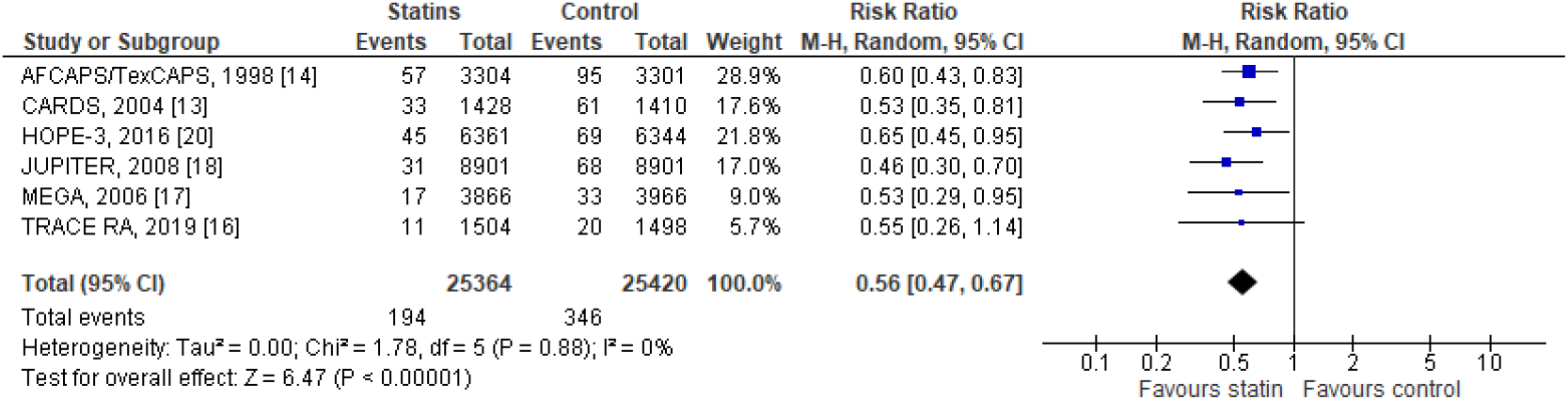
The occurrence of myocardial infarction in included trials. MI indicates myocardial infarction.

#### (2) Major Cerebrovascular Events

This composite outcome that accounted for any stroke, fatal or nonfatal stroke. Nine trials [11,13-20] with 57754 participants, representing 98.7% of the total population, reported stroke. AFCAPS/TexCAPS [14] reported fatal and nonfatal stroke, ALLHAT-LLT [15] reported fatal and nonfatal stroke, CARDS [13] reported strokes, HOPE-3 [20] reported stroke, JUPITER [18] reported any stroke, MEGA [17] reported stroke, PREVEND IT [11] reported hospitalization for cerebrovascular accident, PROSPER reported fatal and non-fatal stroke, and TRACE RA [16] reported presumed ischemic stroke. During the follow-up period, 329/28849 (1.14%) developed stroke in the statin group compared with 419/28905 (1.45%) in the control group. Participants in the statin group had a significantly lower occurrence of stroke in comparison with those of the control group (RR = 0.78, 95% CI: 0.63 to 0.96, I^2^ = 47 %), as can be seen in Figure 4.

**Figure 4:**
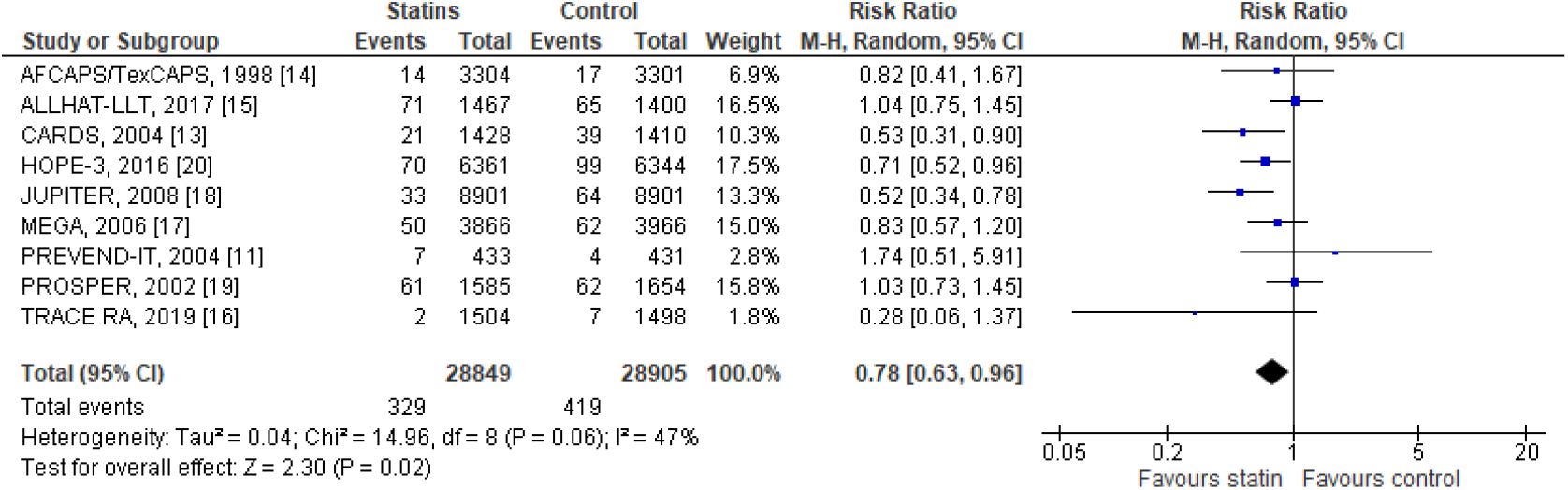
The occurrence of major cerebrovascular events in the included trials.

#### (3) Major Coronary Events

This outcome was defined as coronary heart disease (CHD) death and nonfatal MI. Eight trials [13-20] with 56890 participants, representing 97.2% of the total population, reported major coronary events. During the observation period, 437/28416 (1.54 %) participants developed major coronary events in the statin group compared with 623/28474 (2.19 %) in the control group. Remarkably, participants in the statin group had a lower occurrence of major coronary events in comparison with those of the control group (RR = 0.67, 95% CI: 0.57 to 0.80, I^2^ = 44%), as can be seen in Supplementary Material 4 (Supplementary Figure 1).

#### (4) Composite Cardiovascular Outcome

This outcome included the primary endpoints of most of the trials included in the meta-analysis. However, Beishuizen et al. [12] and HYRIM [10] had non-clinical outcomes as their primary endpoints. ALLHAT-LLT [15] primary preventive analysis did not report outcomes as primary and secondary outcomes. We included acute coronary events in the composite outcome for CARDS [13], as the trial reported a subgroup analysis based on age for this outcome. Eleven trials [10-20] with 58504 participants reported a composite cardiovascular outcome. Significant heterogeneity (I^2^ = 55%) was seen in this outcome, so sensitivity analysis was done for this outcome. During the follow-up period, 956/29235 (3.27%) participants developed composite cardiovascular outcomes in the statin group compared with 1331/29269 (4.55%) in the control group. A remarkable difference existed in both groups, and statins exhibited an apparent decrease in the occurrence of composite cardiovascular outcome (RR = 0.71, 95% CI: 0.62 to 0.82), as can be seen in Supplementary Material 4 (Supplementary Figure 2).

#### (5) CHD Mortality

This outcome was defined as fatal MI, other acute CHD deaths, coronary deaths, or fatal CHD events. Five trials [13-17] with 23144 participants, representing 39.6% of the total population, reported CHD deaths. During the study period, 84/1156 (0.73%) died due to coronary heart disease compared to 95/11575 (0.82%) in the control group. The meta-analysis revealed that there was no statistically significant difference in CHD mortality between the statin and control groups (RR = 0.86, 95% CI: 0.64 to 1.15, I^2^ =0%), as can be seen in Supplementary Material 4 (Supplementary Figure 3).

#### (6) Cardiovascular Mortality

Eight trials [11,13-18,20] with 54515 participants, representing 93.2% of the total population, reported deaths due to cardiovascular causes. The trials included in this outcome had follow-ups ranging from a median of 1.9 years (JUPITER) [18] to a median of 5.6 years (HOPE-3 trial) [20]. During the study period, 341/27264 (1.25%) died due to cardiovascular causes in the statin group compared with 377/27251 (1.38%) in the control group. There was no statistically significant difference in cardiovascular death between the statin and control groups (RR = 0.90, 95% CI: 0.78 to 1.04, I^2^ = 0%), as can be seen in Figure 5.

**Figure 5:**
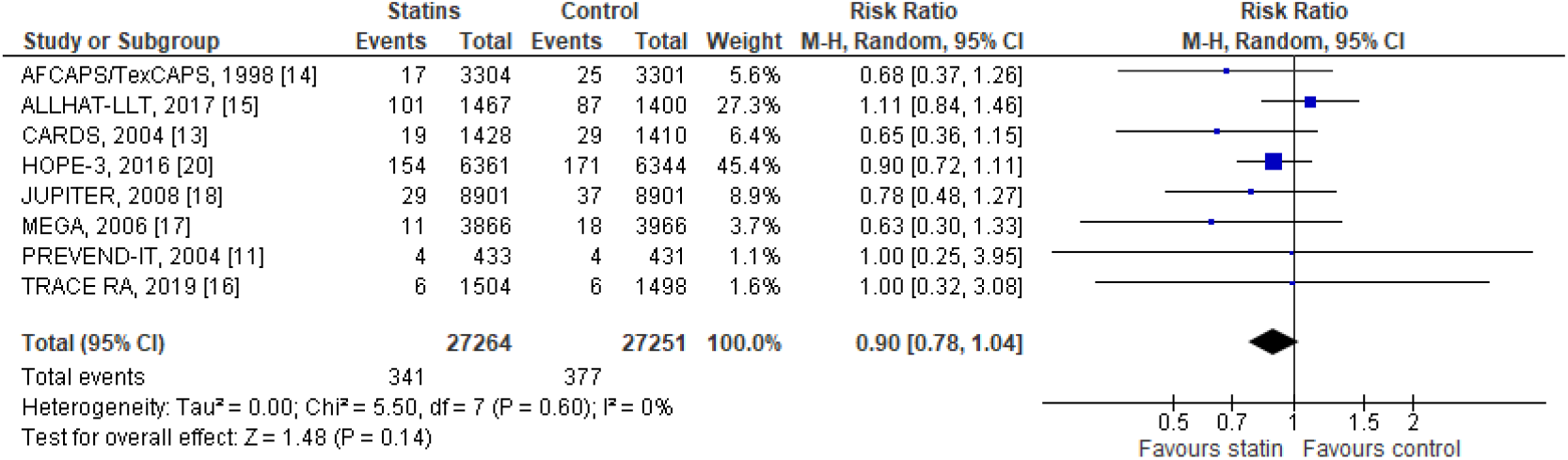
The occurrence of cardiovascular mortality in the included trials.

#### (7) All-cause Mortality

Eleven trials [10-20] with 58504 participants reported all-cause mortality. The duration of follow-up ranged from a median of 1.9 years to a median of 5.6 years. During the observation period, 1169/29235 (3.99%) died in the statin group compared with 1259/29269 (4.30%) in the control group. The meta-analysis did not reveal any statistically significant difference in all-cause mortality between the statin and control groups (RR = 0.92, 95% CI: 0.83 to 1.02, I^2^ =25%), as can be seen in Figure 6.

**Figure 6:**
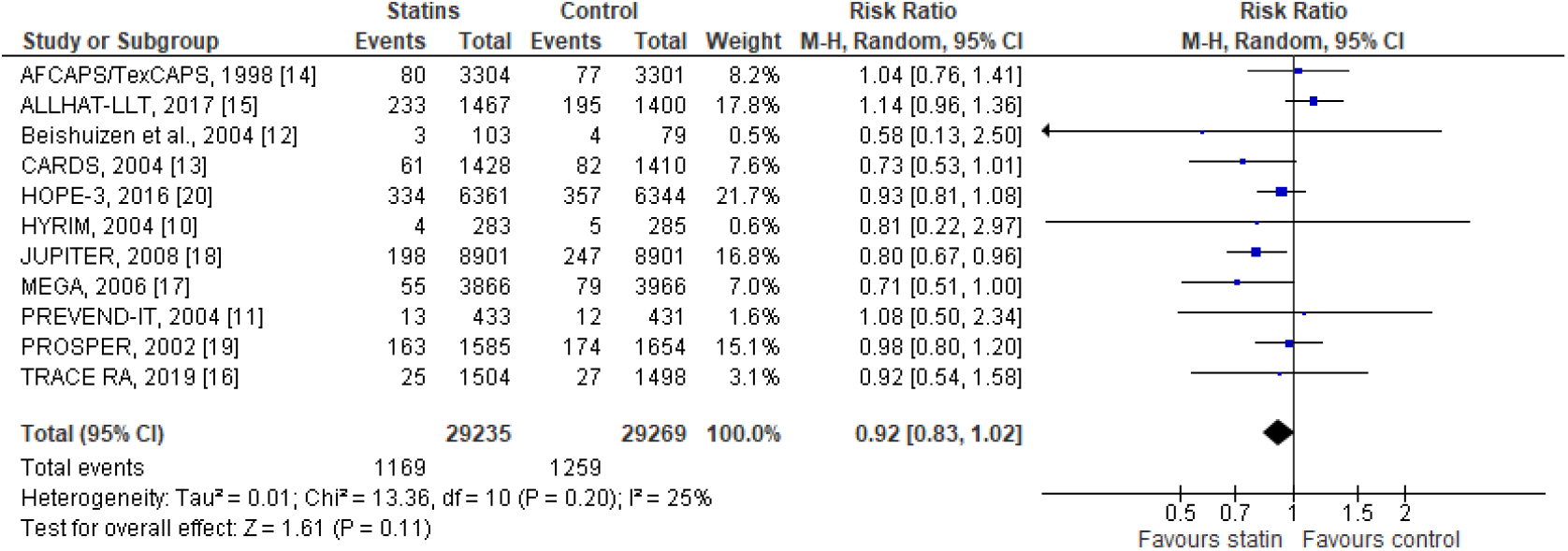
The occurrence of all-cause mortality in the included trials.

#### (8) Muscle-related Adverse Events

Six trials [12-14,16,18,20] with 43134 participants representing 73.7% of the total population, reported myalgia. Four trials [10,13,14,17] with 17264 participants, representing 29.5% of the total population, reported creatine kinase (CK) elevation 10 times the upper limit of normal. Three trials [14,18,20] with 37112 participants, representing 63.4% of the total population, reported rhabdomyolysis. Three trials [13,18,20] with 33365 participants, representing 57% of the total population, reported myopathy (muscle symptoms + CK > 10 times the upper limit of normal). As shown in Supplementary Material 4 (Supplementary Figure 4), there were no statistically significant differences in the muscle-related adverse events between the statin and control groups.

#### (9) Incidence of any Cancer

Eight trials [12-18,20] with 53830 participants, representing 92% of the total population, reported the occurrence of any cancer among the participants. 1168/26931 (4.34%) participants in the statin group reported cancer compared with 1204/26899 (4.48%) in the control group. The meta-analysis did not reveal any statistically significant difference in the cancer incidence between the statin and control groups (RR = 0.97, 95% CI: 0.89 to 1.05, I^2^ = 0%), as shown in Supplementary Material 4 (Supplementary Figure 5).

#### (10) Incident Diabetes

Four trials [14,17,18,20] with 42804 participants, representing 73.2% of the total population, reported the incidence of diabetes mellitus among the participants. 746/21369 (3.49%) participants in the statin group reported diabetes compared with 680/21435 (3.17%) participants in the control group. The meta-analysis did not show any statistically significant difference between the statin and control groups in the occurrence of diabetes mellitus (RR = 1.10, 95% CI: 0.99 to 1.22, I^2^ = 6%), as can be seen in Supplementary Material 4 (Supplementary Figure 5).

### 3.5 Secondary Outcomes

#### (1) Revascularizations

This outcome is comprised of revascularizations, coronary revascularizations, or arterial revascularization. Seven trials [13,14,16-20] with 54023 participants, representing 92.3% of the total population, reported revascularizations. During the follow-up period, 342/26949 (1.27%) participants underwent revascularization procedures in the statin group compared to 533/27074 (1.97%) participants in the control group. Participants in the statin group had a significantly lower occurrence of revascularization procedures compared with those in the control group (RR = 0.65, 95% CI: 0.57 to 0.74, I^2^ = 0%), as can be seen in Supplementary Material 4 (Supplementary Figure 6).

#### (2) Angina

This is the composite outcome that included unstable angina, hospitalization for unstable angina, or angina. Five trials [13,14,17,18,20] with 47782 participants, representing 81.7% of the total population, reported angina. During the study period, 185/23860 (0.78%) participants in the statin group developed angina compared with 244/23922 (1.02%) in the control group. Participants in the statin group had a significantly lower incidence of angina compared with those in the control group (RR = 0.76, 95% CI: 0.63 to 0.92, I^2^ = 0%), as can be seen in Supplementary Material 4 (Supplementary Figure 7).

#### (3) Hospitalization for Cardiovascular Causes

Three trials [11,18,20] with 31371 participants, representing 53.6% of the total population, reported data for hospitalization due to cardiovascular causes. During the study period, 305/15695 (1.94%) participants in the statin group were hospitalized for cardiovascular causes compared with 411/15676 (2.62%) in the control group. Remarkably, participants in the statin group had significantly lower hospitalization compared with those in the control group (RR = 0.74, 95% CI: 0.64 to 0.86, I^2^ = 0%), as can be seen in Supplementary Material 4 (Supplementary Figure 8).

#### (4) Withdrawal of Drug due to Adverse Events

Six trials [11,13,14,16,17,20] with 33846 participants, representing 57.9% of the total population, reported withdrawal of the drug due to adverse events. 1417/16896 (8.39%) participants in the statin group withdrew the drug compared with 1521/16950 (8.97%) participants in the control group. There was no statistically significant difference between the statin and control groups in the withdrawal of the drug due to adverse events, but significant heterogeneity was present (RR = 0.91, 95% CI: 0.69 to 1.18, I^2^ = 90%), as can be seen in Supplementary Material 4 (Supplementary Figure 5).

#### (5) Serious Adverse Events

Five trials [13,14,16,18,20] with 42952 participants, representing 73.4% of the total population, reported serious adverse events. 2615/21498 (12.16%) participants had serious adverse events in the statin group compared with 2639/21454 (12.30%) participants in the control group. The meta-analysis did not reveal any difference in serious adverse events between the statin and control groups (RR = 0.99, 95% CI: 0.95 to 1.04, I^2^ = 0%), as can be seen in Supplementary Material 4 (Supplementary Figure 5).

#### (6) Fatal Cancer

Four trials [13,14,18,20] with 39950 participants, representing 68.3% of the total population, reported fatal cancer. During the observation period, 211/19994 (1.06%) participants had fatal cancer in the statin group compared with 236/19956 (1.18%) in the control group. However, there was no statistically significant difference in fatal cancer between the statin and control groups (RR = 0.87, 95% CI: 0.61 to 1.23, I^2^ = 66%), as can be seen in Supplementary Material 4 (Supplementary Figure 5).

#### (7) Liver Enzyme Abnormalities

It was defined as AST/ALT elevation two times the upper limit of normal. Six trials [12-14,16-18] with 38146 participants, representing 65.2% of the total population, reported liver enzyme abnormalities. 256/19044 (1.34%) participants in the statin group had liver enzyme abnormalities compared with 215/19102 (1.13%) participants in the control group. The meta-analysis did not reveal any statistically significant difference in liver enzyme abnormalities between the statin and control groups (RR = 1.20, 95% CI: 1 to 1.43, I^2^ = 0%), as can be seen in Supplementary Material 4 (Supplementary Figure 5).

### 3.6 Sensitivity Analyses

We did a sensitivity analysis based on no. of participants (> 3000 and < 3000), duration of follow-up (> 3.5 years and < 3.5 years), randomization status in the study (adequate and inadequate), the blinding status of participants (blinded and unblinded) and mean LDL-C of participants (near-optimal and borderline-high range), as shown in Supplementary Material 3 (Supplementary Table 3). We found the estimate of effect size to be homogenous across all these groups except for the outcomes, major cerebrovascular events, and all-cause mortality. The effect size of these outcomes was remarkably significant for the group of trials where the mean LDL-C of participants was in the near-optimal range (100 -129 mg/dl). We created a group that included trials or subgroups of trials where all participants had LDL-C 160 mg/dl and calculated the pooled estimate of risk ratio (RR). We found the risk ratio to be statistically significant for the following outcomes: composite cardiovascular outcome, MI, major cerebrovascular events, major coronary events, all-cause mortality, and revascularizations.

We created a group to include trials or subgroups of trials where all participants had diabetes and calculated the pooled estimate of risk difference (RD), as shown in Supplementary Material 3 (Supplementary Table 4). MI, major cerebrovascular events, major coronary events, and all-cause mortality were the outcomes that had statistically significant risk differences. We then compared the point of estimate of RD for these outcomes with the point of estimate of RD for the corresponding outcomes of all trials and found a greater effect in the diabetic group.

Among the different systemic inflammation mediators, C-reactive protein (CRP) has been widely accepted as a potential independent risk indicator of future cardiovascular events [21]. Hence, we created a group that included JUPITER [18] (where all the participants had high-sensitivity CRP ≥2 mg/L), the HOPE-3 trial [20] subgroup (where all the participants had high-sensitivity CRP > 2mg/L) [20], and TRACE RA [16] (where all the participants had inflammatory condition Rheumatoid arthritis), as shown in Supplementary Material 3 (Supplementary Table 4). Therefore, all participants in this group were at an increased risk of CVD. We calculated the pooled estimate of RD for this group, and the effect size was significant for the following outcomes: composite cardiovascular outcome, MI, major cerebrovascular events, major coronary events, all-cause mortality, and revascularizations. On comparing the point of estimate of RD for these outcomes with the point of estimate of RD for the corresponding outcomes of all trials, we found a superior effect estimate for major cerebrovascular events and all-cause mortality.

### 3.7 Publication Bias

We assessed publication bias by constructing a funnel plot for the outcomes that included ten or more trials. Therefore, we created a funnel plot for the composite cardiovascular outcome and all-cause mortality. We checked for publication by examining the shape of the funnel plot for any asymmetry. Supplementary Material 4 (Supplement Figure 9) shows the funnel plot for the composite cardiovascular outcome, and Figure 7 shows the funnel plot for all-cause mortality.

**Figure 7:**
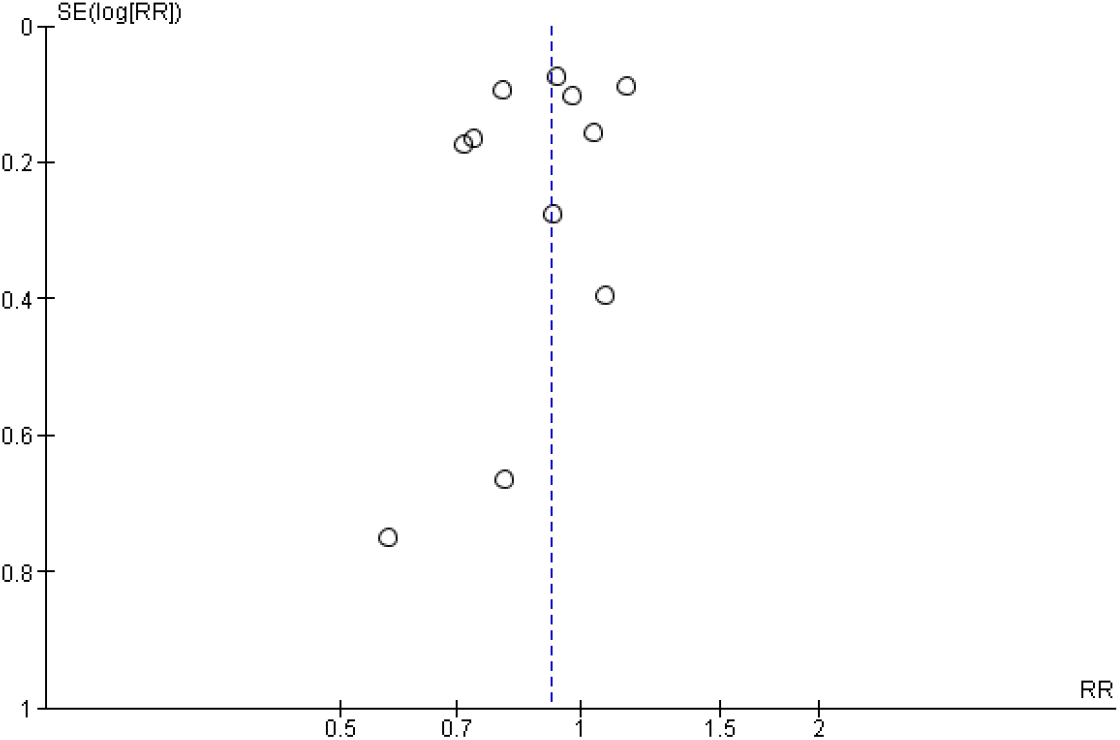
Funnel plot for all-cause mortality.

### 3.8 GRADE of Evidence

We assessed the quality of evidence for important outcomes via the GRADE approach using GRADEpro GDT software. Most outcomes had moderate scores. Others had high scores, and one outcome had a low score. Inconsistency, lack of reporting of outcome in some trials, and an imprecise estimate of effect size were the reasons for moderate and low scoring. The GRADE quality of summary evidence for important outcomes is shown in Supplementary Material 3 (Supplementary Table 5).

## 4. Discussion

We performed a meta-analysis of 11 randomized controlled trials to find evidence on the role of statins in primary prevention of atherosclerotic cardiovascular disease and mortality, and harms in a population whose baseline average LDL-C ranged from near-optimal to borderline high levels in standard practice. Meta-analyses of statin trials have shown benefits for mortality, cardiovascular diseases, and procedures [22,23]. Existing guidelines adapted to the evidence from the included trials and recommended statins in the primary prevention of cardiovascular disease, not limiting statins as a lipid-lowering agent [24]. Primary prevention trials and meta-analyses provide evidence in favor of statins [25-30]. However, existing meta-analyses included trials where greater than 10% of the population had cardiovascular disease or subclinical atherosclerosis or had a baseline mean LDL-C from near-optimal to high levels [25-30]. A meta-analysis that included both primary and secondary prevention trials showed that total and cardiovascular mortality benefit in baseline LDL-C greater than 100 mg/dl [22]. A knowledge gap exists on the primary preventive role of statins in populations with baseline average LDL-C in near-optimal to borderline high range. We included a new trial [16] to find up-to-date evidence; analyzed trials where near all populations are free from prior cardiovascular diseases; and with a mean of baseline LDL-C less than 160, among which, two trials and a subgroup of a trial included all participants with LDL-C less than/equal to 160 mg/dl.

We found a reduction in cardiovascular diseases and revascularization procedures without an increase in adverse events. Our findings are consistent with other meta-analyses, except for Chen et al. [25], who did not find statin therapy to reduce coronary revascularizations. Statin therapy did not reduce all-cause mortality, cardiovascular mortality, and CHD mortality. Only the JUPITER trial [18] found a significant risk reduction in all-cause mortality. Other meta-analyses had inconsistent evidence regarding all-cause mortality; Chou et al. [26], Brugts et al. [30], and Taylor et al. [28] reported in favor of statin therapy, whereas Chen et al. [25], Thavendiranathan et al. [29], and Li et al. [27] reported no significant effect. Chou et al. [26] and Thavendiranathan et al. [29] found statins reduced cardiovascular mortality, but Li et al. [27] found inconsistent evidence, where the significant reduction seen in the fixed-effect analysis was lost in random-effects analysis. We suggest no benefit of statin therapy on reducing cardiovascular mortality in a population with baseline average LDL-C in near-optimal to borderline high range, which is consistent with nine trials that reported the outcome. Statins did not reduce CHD mortality, which is supported by six trials that reported the outcome and meta-analysis by Thavendiranathan et al. [29] and Li et al. [27].

The JUPITER trial [18] showed an increased risk of type II diabetes mellitus and decreased risk of fatal cancer. The HOPE-3 trial [20] showed an increased risk of myalgia, but Beishuizen et al. [12] showed a decreased risk. The CARDS trial [13] showed a decreased risk of creatine kinase elevation. We found no risk of any adverse events with statin therapy, with heterogeneity seen in myalgia, liver enzyme abnormalities, or fatal cancer. Other meta-analyses [25,26,29,30] support our findings, whereas Taylor et al. [28] and Li et al. [27] oppose our findings and support the JUPITER trial [18] that found an increased risk of diabetes mellitus. Withdrawal due to adverse events was not associated with statin therapy, but significant heterogeneity was present. The MEGA trial [17] found increased withdrawal in the statin group, and the HOPE-3 trial [20] found increased withdrawal in the control group. Our finding is consistent with the findings of Chou et al. [26].

In the sensitivity analysis, statin therapy remained consistent in reducing composite cardiovascular outcomes and myocardial infarction irrespective of participant’s number, quality of trials (randomization or blinding), baseline mean LDL-C, and duration of statin therapy. However, no. of participants was determining for few outcomes like major cerebrovascular events, revascularizations, and angina, where trials with less than 3000 participants did not show any benefit, which remained consistent in pooled estimates of those trials. Quality of trials was important for few outcomes because trials with moderate quality (unclear randomization and unblinding) could not demonstrate a reduction in major cerebrovascular events, major coronary events, and angina, but our pooled estimate showed clear benefit. Shorter trials (less than 3.5 years) failed to demonstrate the benefit of statins in major cerebrovascular events, major coronary events, and angina, but our pooled estimate showed a clear benefit. However, the benefits evident in reducing all-cause mortality in larger trials (> 3000 participants) and good quality (blinding) questions our pooled estimate that did not find a benefit. The JUPITER [18], a successful primary prevention trial, and meta-analyses by Chou et al. [26], Brugts et al. [30], and Taylor et al. [28] also oppose our findings and provide evidence of all-cause mortality reduction. Thus, the benefit of statins in reducing all-cause mortality remains inconsistent and warrants larger (> 3000) and high-quality trials to find better evidence. CHD mortality reduction was shown in the ALLHAT-LLT trial [15] and MEGA trial [17], but both trials were unblinded, so the benefit is still questionable because our meta-analysis did not find it to be true.

Our analysis of trials or subgroups of trials that included all participants with diabetes mellitus was consistent with our overall pooled estimate, but the risk reduction in MI, major cardiovascular events, and major coronary events was superior in diabetic populations. In addition, all-cause mortality reduction was evident in the diabetic population that was not present in our overall meta-analysis of the included trials. Thus, all-cause mortality reduction is evident with statin therapy in the diabetic population. As opposed to our findings, the benefit of composite cardiovascular outcome was not seen in the diabetic population, but significant heterogeneity was present, making the finding questionable. The lack of benefit of revascularizations and angina in the diabetic population could be due to only CARDS [13], contributing to the finding, which lacked power because of only 2838 participants with 58 and 16 events in revascularizations and angina, respectively; our pooled estimate of trials with participants below 3000 also could not find significance in revascularization and angina reduction. The benefit of statin therapy in reducing MI evident in our meta-analysis is consistent in the diabetic population, which is considered a clear ASCVD risk factor by current guidelines (3). The benefit of reducing CHD mortality in the diabetic population opposed our findings and showed that statin has a benefit in CHD mortality in the diabetic population.

In our analysis of trials or subgroup of trials where all participants had increased risk of CVD based on high-sensitivity C-reactive protein (hs-CRP) ≥2 mg/L (JUPITER [18]and the subgroup of HOPE-3 trial [20]) and participants having Rheumatoid arthritis (TRACE RA [16]), the findings are consistent to our meta-analysis findings with superior risk reduction compared to overall population with an additional benefit in reducing all-cause mortality, except benefit in reducing angina was lost, for which only JUPITER [18] contributed, which terminated early with median 1.9 years; it is consistent with our finding that shorter trials (< 3.5 years follow-up) failed to show benefit in angina reduction. Hs-CRP is acknowledged as an ASCVD risk factor in current guidelines, and our findings of benefit in this population are consistent with the guidelines [24].

In our analysis of trials or subgroups of trials with all participants having LDL-C ≤ 160 mg/dl, our findings are consistent and with superior risk reduction in this population. An additional benefit of all-cause mortality is present in this population as opposed to our findings of the overall analysis. The loss of angina benefit is probably due to the CARDS trial [13] having less than 3000 population and JUPITER trial [18] having < 3.5-year follow-up, which is consistent with our analysis of angina being affected by participant number in trials (below 3000) and study duration (less than 3.5 years). People of south Asian ancestry is already acknowledged as an ASCVD risk enhancer, and we found the benefit of statin therapy in participants with baseline LDL-C ≤ 160 mg/dl. The current guidelines recommend statin therapy after assessment of 10-year ASCVD risk in the south Asian ethnic group with LDL-C ≥70 mg/dl and < 190 mg/dl. The recommendation needs consideration for this ethnic group because the population in South-Asian regions has mean cholesterol lower than western standards [5], so they might be deprived of the benefits of statins, considering the evidence of the cardiovascular benefit of statins in the LDL-C range 100 to 159 mg/dl in our meta-analysis.

Mean cholesterol among different populations differs significantly, and of many factors, diet is the major one. Populations where there is high consumption of saturated fat and lower consumption of polyunsaturated fat, have higher cholesterol levels [1]. Since the relationship between blood cholesterol level and CVD risk appears to be continuous without any threshold, if the threshold for “high cholesterol” is set at over 147 mg/dl, then this could lead to 4.4 million deaths and 40.4 million disability-adjusted life years (DALYs) worldwide [1].

Cholesterol Treatment Trialists’ (CTT) collaboration 2010 meta-analysis demonstrated that each 1 mmol/L reduction in LDL-C reduces the annual rate of major vascular events by just over a fifth, and LDL-C reduction by 2-3 mmol/L would further reduce the risk by about 40-50% [23]. Hence, CTT Collaboration 2010 meta-analysis established LDL-C to be an important risk factor for major vascular events [23]. The 2019 American College of Cardiology and American Heart Association (ACC/AHA) guideline has lipid-based criteria to initiate statin therapy if LDL-C ≥190 mg/dl [24]. Still, our meta-analysis found evidence of benefit with no increase in the risk of harm in the population with LDL-C lower than 190 mg/dl and some cardiovascular risk factors. Population with average LDL-C from near-optimal to borderline high is not getting the benefit of statin therapy according to the guidelines when 10-year ASCVD risk criteria are not met, which might be important in the south Asian population who is already accepted to have enhanced ASCVD risk and a population whose average LDL-C is low compared to western standards [5]. In our meta-analysis, we found overall cardiovascular benefit in treating the population with baseline average LDL-C in near-optimal to borderline high range, and more benefit appears in those having traditional risk factors of CVD. Hence, further research seems necessary to analyze cost-effectiveness and risk-benefit ratio to consider starting statin therapy in a population with mean baseline LDL-C in near-optimal to borderline high range and low ASCVD risk according to current guidelines [24]. Also, the 2019 ACC/AHA guidelines have set the criteria to initiate statin therapy in the age group 20-39 year if a family history of premature ASCVD and LDL-C ≥160 mg/dl are both present [24]. However, our meta-analysis showed the overall cardiovascular benefit in treating the LDL-C range of 100-159 mg/dl, and following this lipid criteria in populations where average cholesterol is lower than the western standard seems to bar these populations from the cardiovascular benefit of statins.

## 5. Limitations

This study has several limitations. We were able to include only trial-level data. Patient-level data would have enabled us to include participants free of prior cardiovascular disease because few trials included participants with prior cardiovascular diseases. This study included trials with a baseline average LDL-C in the study population less than 160 mg/dl, so participants with even higher baseline LDL-C might have contributed to the findings, but we did sensitivity analysis of trials and subgroups of trials with all participants having LDL-C ≤ 160 mg/dl. This study included participants with heterogeneous cardiovascular risk factors (hypertension, diabetes, elevated hs-CRP), which might have contributed to the heterogeneous effect size, but significant heterogeneity was seen in outcomes: composite cardiovascular outcomes, myalgia, liver enzyme abnormality, withdrawal due to adverse events, and fatal cancer. We did a sensitivity analysis based on participants and study characteristics to check for uniformity in the effect size in different participant cohorts and quality of studies. Statin therapy in the trials varied in statin type, dosage, or mode of therapy (fixed or titrated dose) with two trials with high-intensity statin [16,18], six with moderate intensity stain [11-13,15,19,20], and two with low [10,17] and one with a low and moderate-intensity statin [14]. Trials in the study included the population with age greater than 18, and the mean age of included participants in all trials was above 40, so this study is limited in finding the benefit in a younger population less than 40 years of age. We included published data in the English language only, which could have resulted in the loss of trials published in other languages. All trials had some form of pharmaceutical sponsorship, which might have led to reporting bias or attrition bias in fewer trials, especially in adverse events of statin therapy.

## 6. Conclusions

In this study, we found the benefit of statin therapy in reducing cardiovascular diseases and procedures with no increased risk of harms in the population with baseline mean LDL-C in near-optimal to borderline-high range without prior atherosclerotic cardiovascular disease. However, we did not find any benefit of statin therapy in reducing cardiovascular and CHD mortality. The all-cause mortality benefit was seen in the population with diabetes and increased risk of cardiovascular disease.

## Data Availability

All supporting data for this meta-analysis are from previously reported randomized controlled trials and meta-analyses, which have been cited.

## Authors’ Contributions

Sanjay S. Srivatsa and Prabhat Adhikari provided guidelines for this systematic review and meta-analysis. Bishnu M. Singh and Hari K. Lamichhane wrote the main manuscript, supplemental material. Sanjay S. Srivatsa, Prabhat Adhikari, Bishnu M. Singh, and Hari K. Lamichhane conducted the literature search, study selection, risk of bias assessment, and quality of evidence assessment via the GRADE approach. Bishnu M. Singh, Hari K. Lamichhane, and Dhan B. Shrestha conducted the data extraction. Hari K. Lamichhane, Bishnu M. Singh, Bikash J. Kshetri, and Sijan Khatiwada conducted the statistical analysis and interpretation of data. All authors actively participated in manuscript preparation and review. Bishnu M. Singh and Hari K. Lamichhane contributed equally to this work.

## Acknowledgments

None.

## Supplementary Materials

There were four supplementary materials. The first one is the “Search Strategies for Different Electronic Databases” the second one is “PRISMA checklist,” and the third one is Supplement Table 1-5: Supplement Table 1: Patient Baseline Characteristics and Interventions Used in the Included Trials. Supplement Table 2: Study Design and Target Population of the Included Trials. Supplement Table 3: Sensitivity Analysis Stratified by the Trial Characteristics. Supplement Table 4: Sensitivity Analysis Stratified for the Type of Population. Supplement Table 5: Grade of Evidence. The fourth Supplementary Material is Supplement Figure 1-9: Supplement Figure 1: Meta-analysis of Major Coronary Events. Supplement Figure 2: Meta-analysis of Composite Cardiovascular Outcome. Supplement Figure 3: Meta-analysis of Coronary Heart Disease Mortality. Supplement Figure 4: Meta-analysis of Muscle-related Adverse Events. Supplement Figure 5: Meta-analysis of Incidence of Other Adverse Events. Supplement Figure 6: Meta-analysis of Revascularizations. Supplement Figure 7: Meta-analysis of Angina. Supplement Figure 8: Meta-analysis of Hospitalizations for Cardiovascular Causes. Supplement Figure 9: Funnel Plot for Composite Cardiovascular Outcomes. (Supplementary Materials)

## Supplemental Material

### Supplementary Material 1

Search strategies for different electronic databases

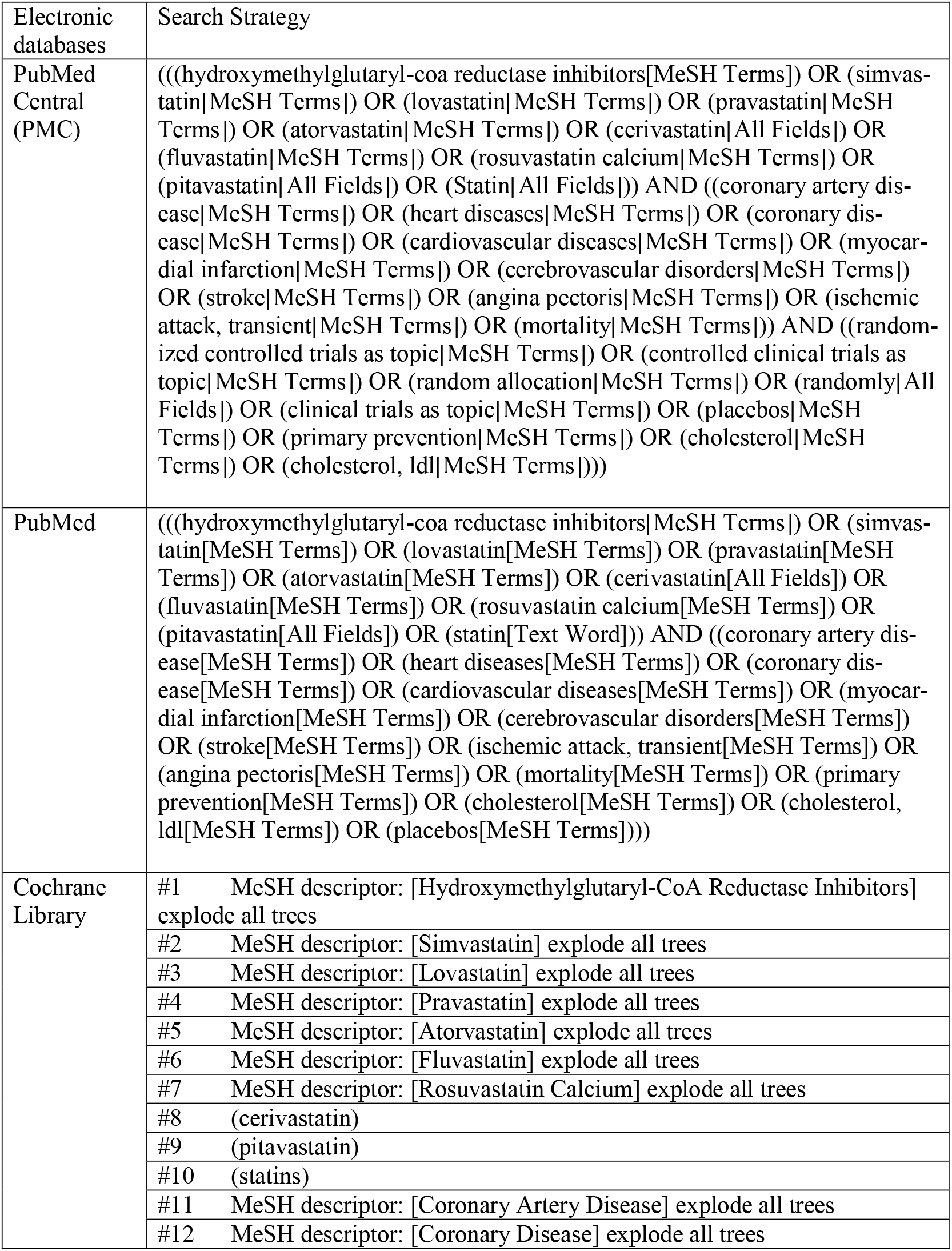
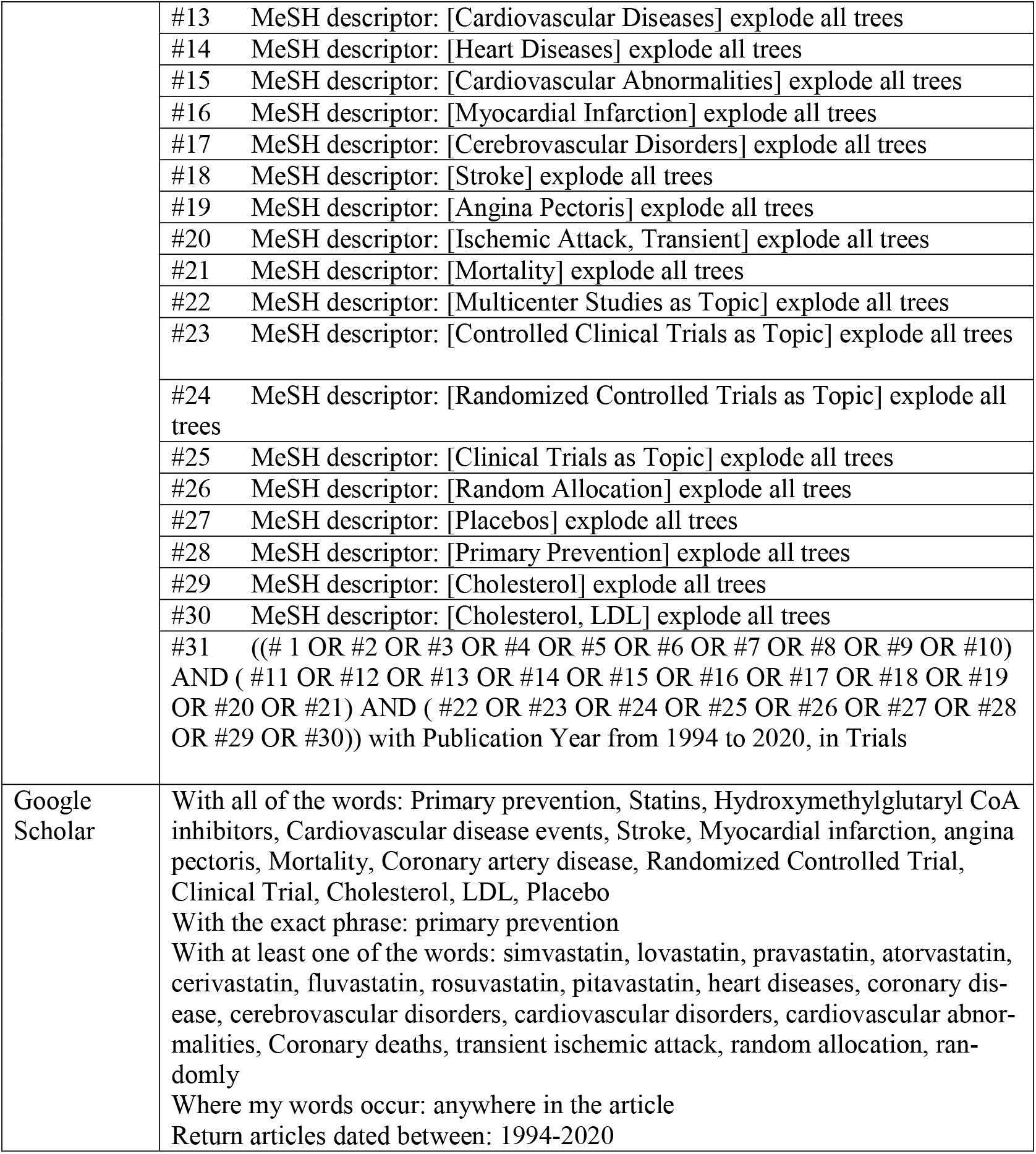

### Supplementary Material 2

PRISMA checklist

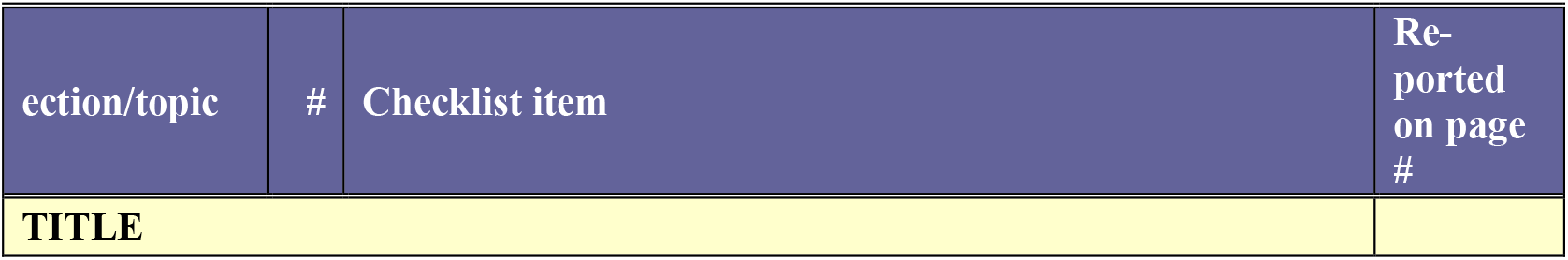
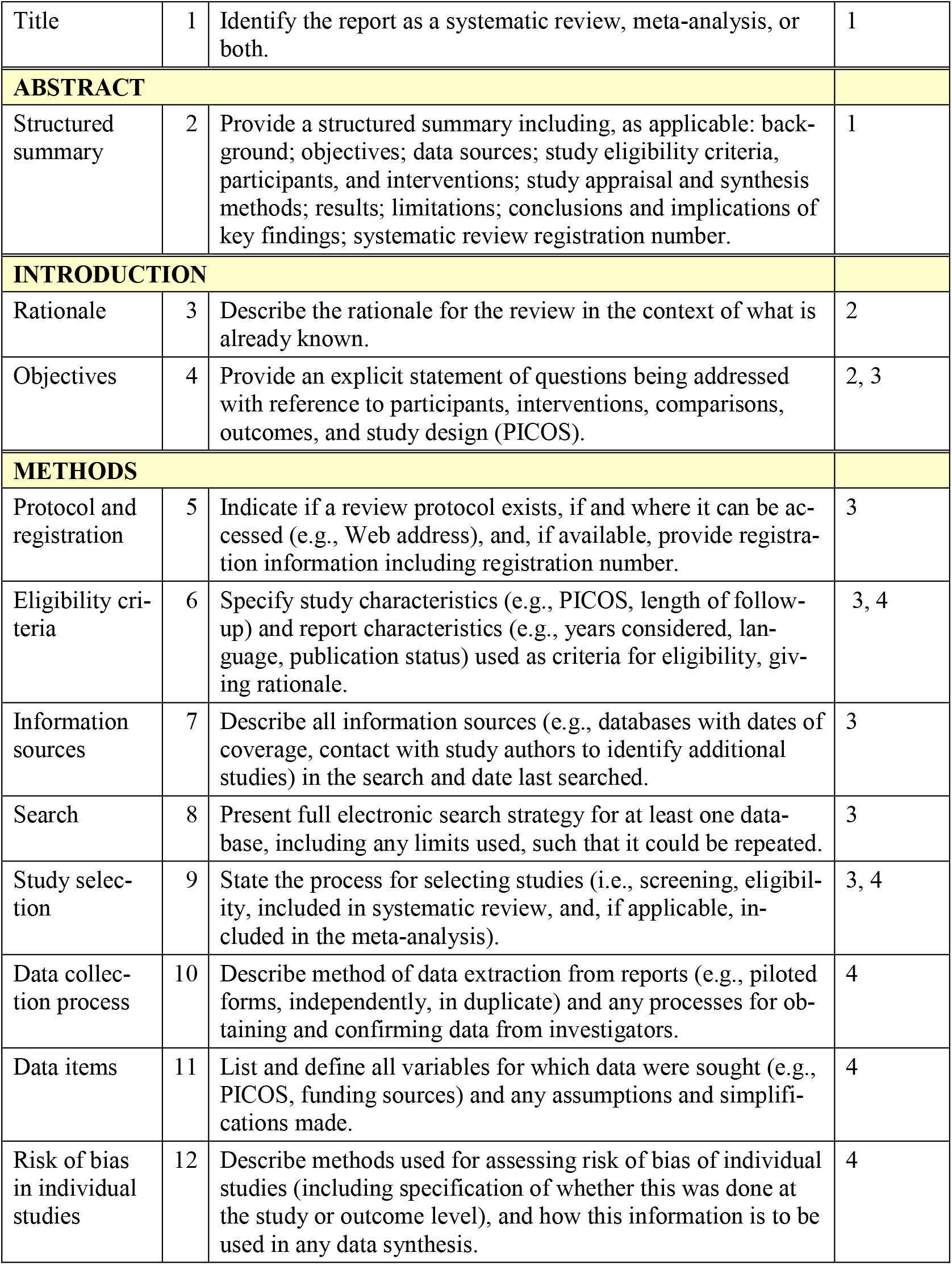
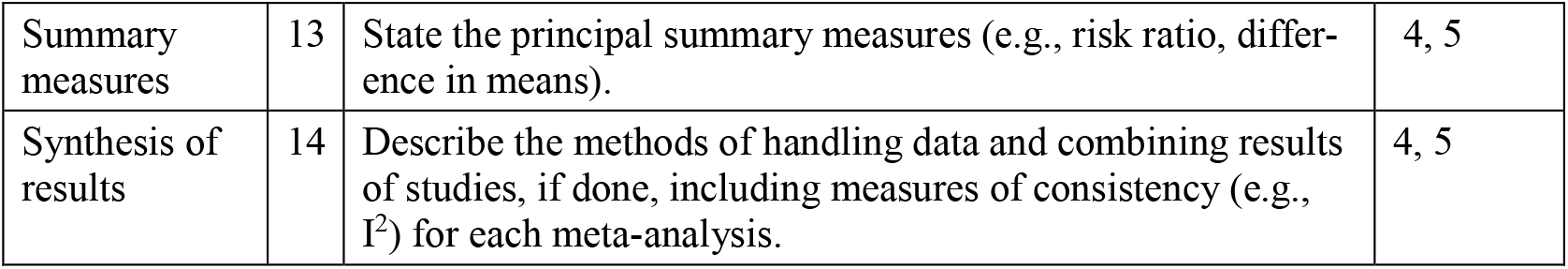

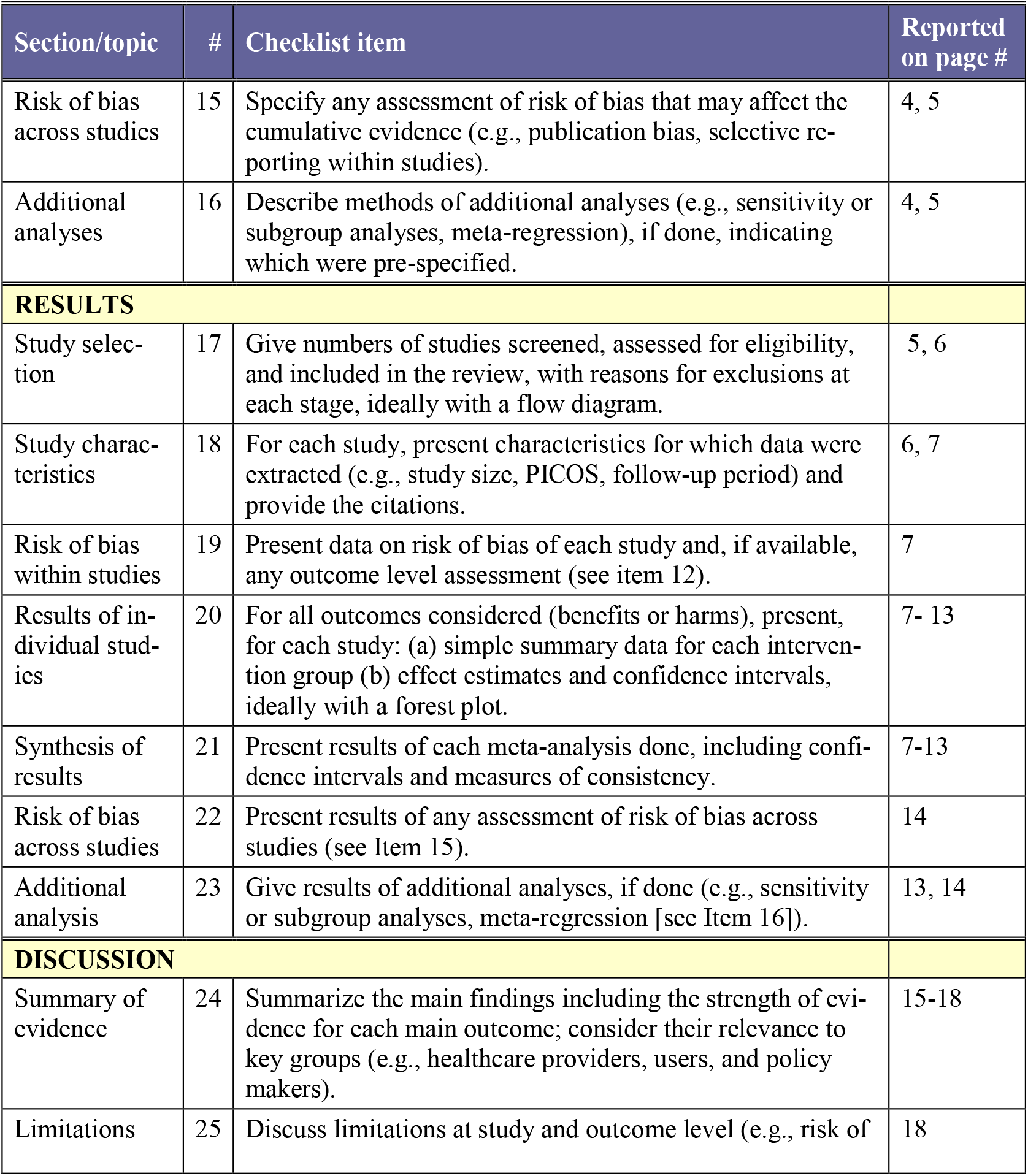
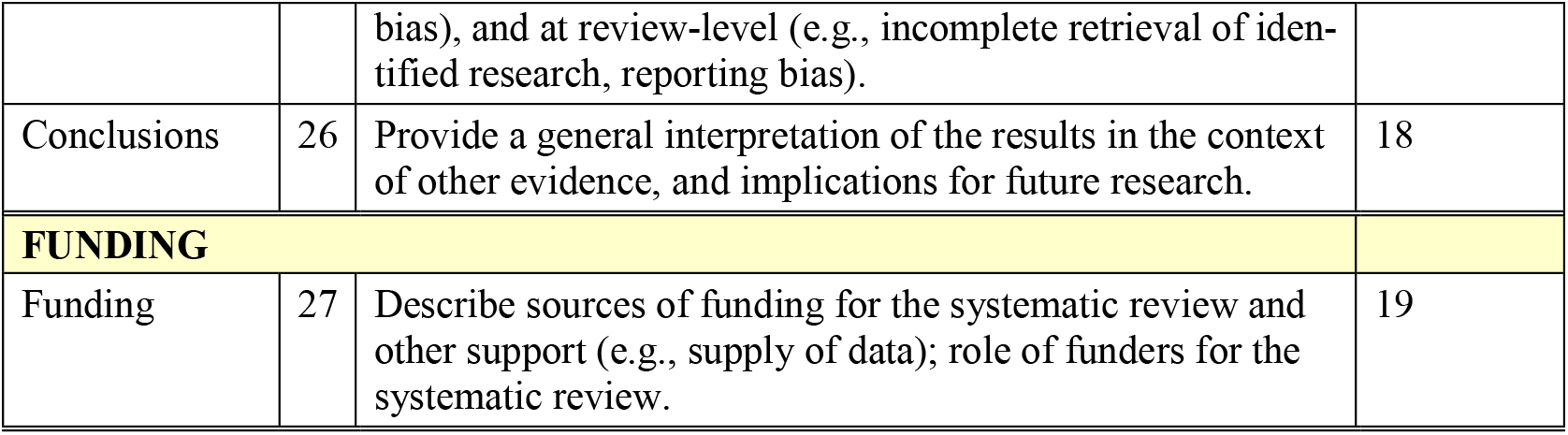

### Supplementary Material 3

Supplement Table 1-5

**Supplement Table 1.**
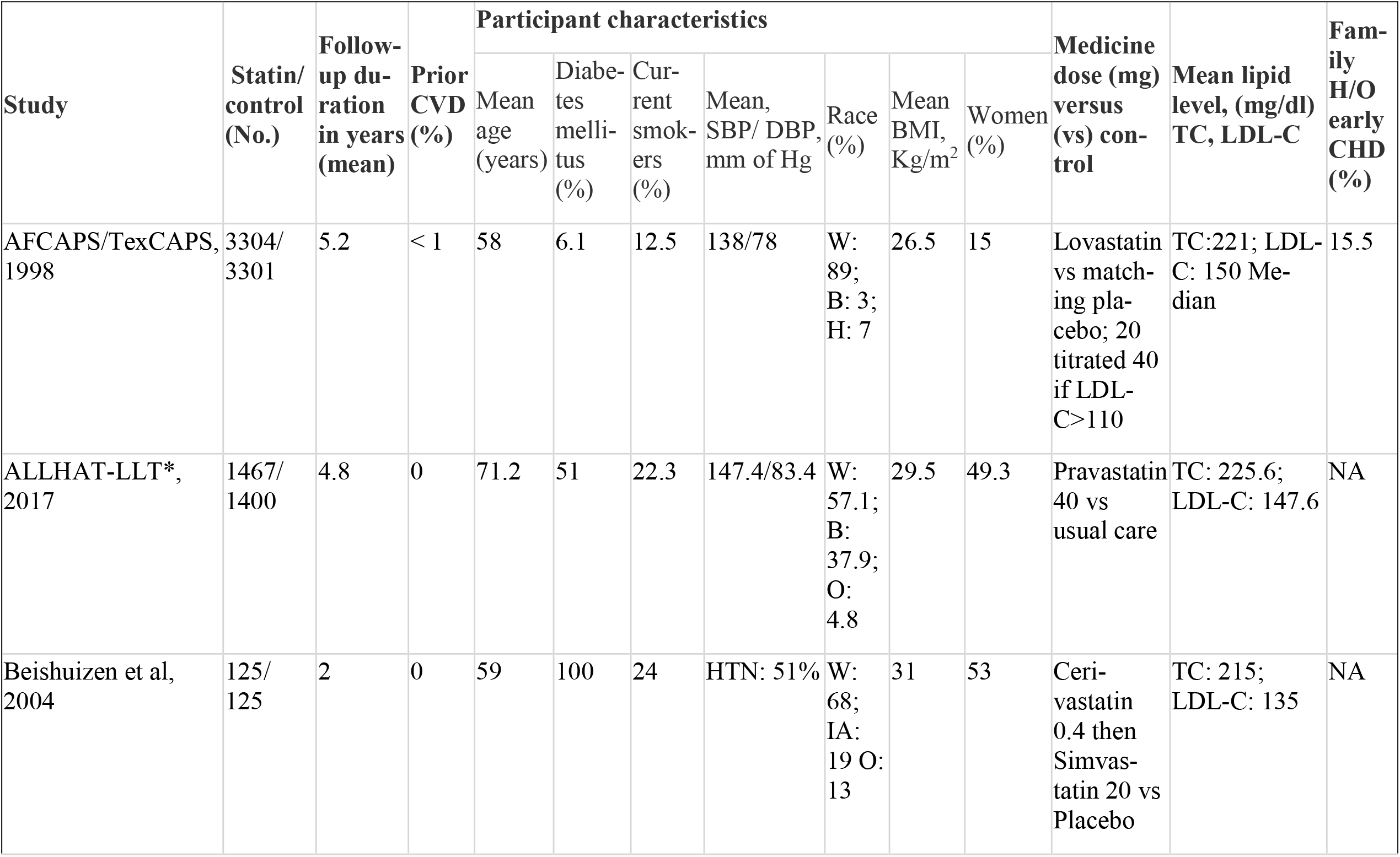
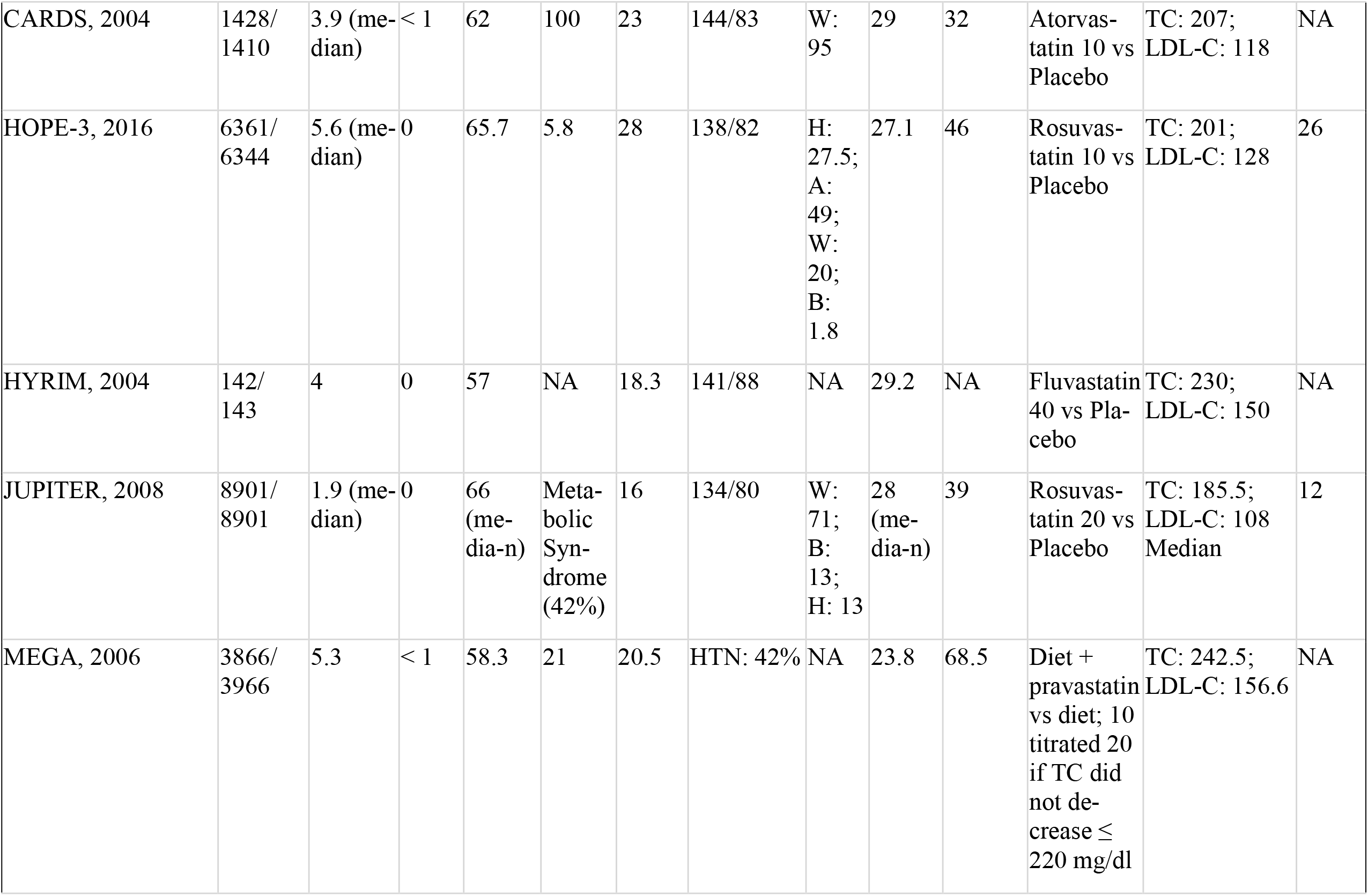
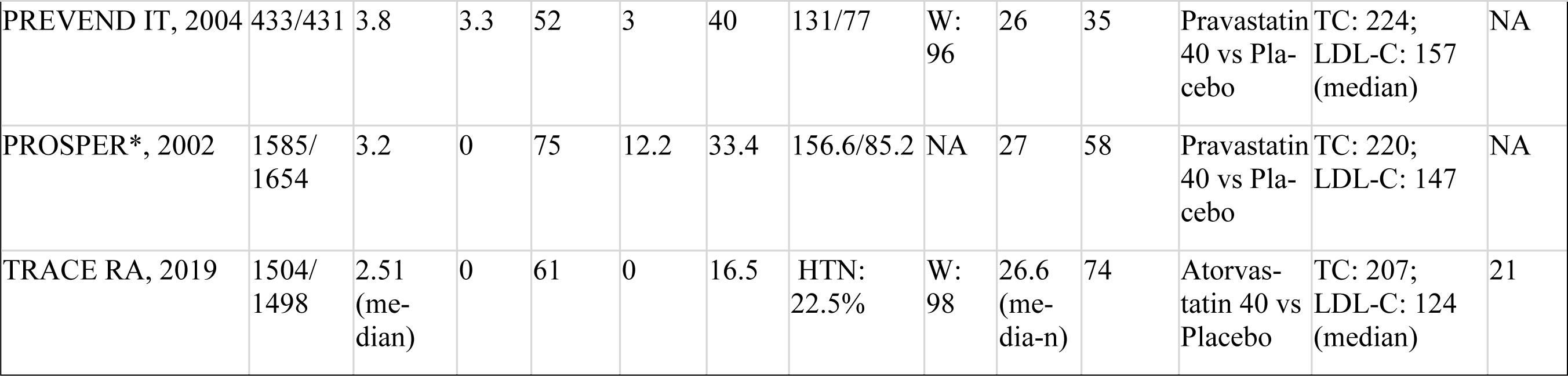
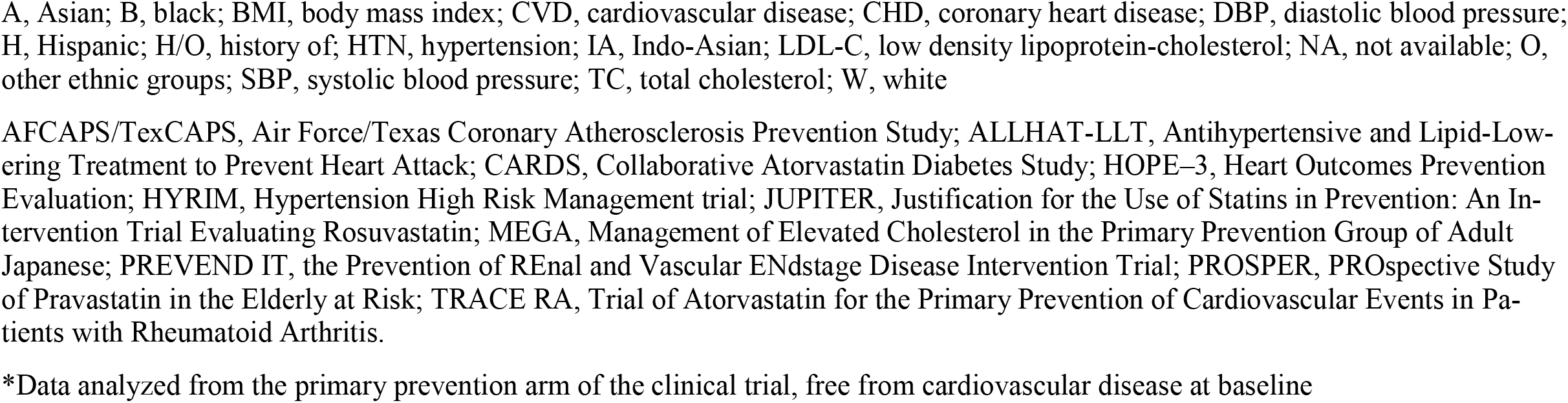
Patient baseline characteristics and interventions used in the included trials

**Supplement Table 2.** Study design and target population of the included trials

| Study | Target Population | Study Design | Statistical Analysis calculation |
| --- | --- | --- | --- |
| AFCAPS/TexCAPS, 1998 | Men and women with average TC and LDL-C and below-average HDL-C without CVD | Randomized, double-blind, placebo-controlled primary prevention trial | Cox proportional hazards regression model |
| ALLHAT-LLT*, 2017 | 2867 older adults $\geq 65$ Y without atherosclerotic cardiovascular disease as a subset of main study ALLHAT-LLT | Randomized, non-blinded, large simple trial | Cox proportional hazards model |
| Beishuizen et al, 2004 | Participants aged 30-80 years without CVD but Type 2 DM for at least one year | Randomized, placebo-controlled, double-blind clinical trial | Clinical events as proportion & evaluate using $\chi^2$ or Fisher's test |
| CARDS, 2004 | Age 40-75 Y with Type 2 DM with at least one: retinopathy, albuminuria, smoker, or HTN. Participants had no CVD, LDL-C $\leq 160$ mg/dl | A multicenter randomized placebo-controlled trial | Cox regression survival analysis |
| HOPE-3, 2016 | Men $\geq 55$ Y and women $\geq 65$ Y without CVD had at least one: elevated waist-to-hip ratio, H/O low HDL-C, current tobacco use, dysglycemia, family H/O premature CHD, and mild renal dysfunction. | A pragmatic, multicenter, long-term, international, double-blind, randomized, placebo-controlled trial based on 2-by-2 factorial design | Cox proportional hazards model |
| HYRIM, 2004 | Men 40-74 Y with HTN, without CVD, and TC 174-309 mg/dl, TG $< 398$ mg/dl, BMI 25-35 kg/m<sup>2</sup>, and sedentary lifestyle. | Randomized, placebo-controlled, 2-by-2 factorial design | Secondary outcome variables as no. of CVD events |
| JUPITER, 2008 | Men ≥ 50 Y and women ≥ 60 Y without CVD and screening LDL-C $<$ 130 mg/dl and high-sensitivity C-reactive protein ≥ 2 mg/L. | Randomized, double-blind, placebo-controlled, multicenter trial | Cox proportional hazards model |
| MEGA, 2006 | Men and postmenopausal women aged 40-70 Y without any H/O CHD or stroke. Participants with familial hypercholesterolemia were excluded. | Prospective randomized, open-labeled, blinded-endpoint study | Cox's proportional hazards model |
| PREVEND IT, 2004 | Participants had persistent microalbuminuria, blood pressure $<$ 160/100 mm of Hg, TC $<$ 309 mg/dl or $<$ 193 mg/dl in case of previous MI | Single-center, double-blind, randomized, placebo-controlled trial with a 2-by-2 factorial design | Two-way ANOVA; HR or RR with 95% CI |
| PROSPER*, 2002 | 3239 participants without cardiovascular disease as a primary preventive subgroup of the main study | Randomized placebo-controlled and blinded trial | HR and P values from the Cox model analyses |
| TRACE RA, 2019 | Rheumatoid arthritis (RA) patients $>$ 50 years of age and had disease $>$10 years duration. Those with CVD, DM, myopathy, taking statins were excluded | Randomized, double-blind, placebo-controlled, multicenter clinical trial | Cox regression models |

ANOVA, analysis of variance; BMI, body mass index; CVD, cardiovascular disease; CI, confidence interval; CHD, coronary heart disease; DM, diabetes mellitus; HR, hazard ratio; HDL-C, high density lipoprotein-cholesterol; H/O, history of; HTN, hypertension; MI, myocardial infarction; RR, risk ratio; TC, total cholesterol; TG, triglyceride; Y, year
\*Data analyzed from the primary prevention arm of the clinical trial, free from cardiovascular disease at baseline

**Supplement Table 3.**
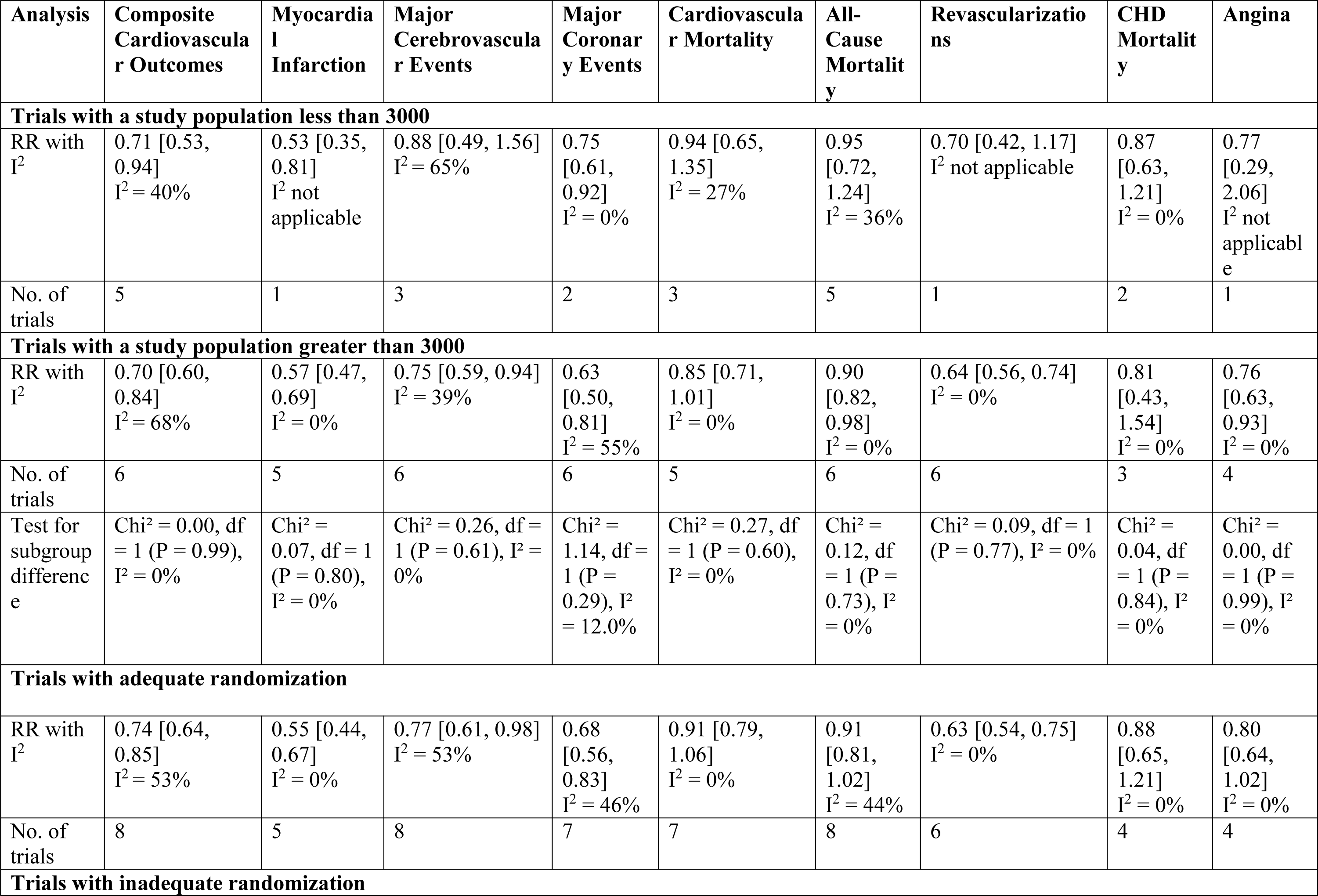
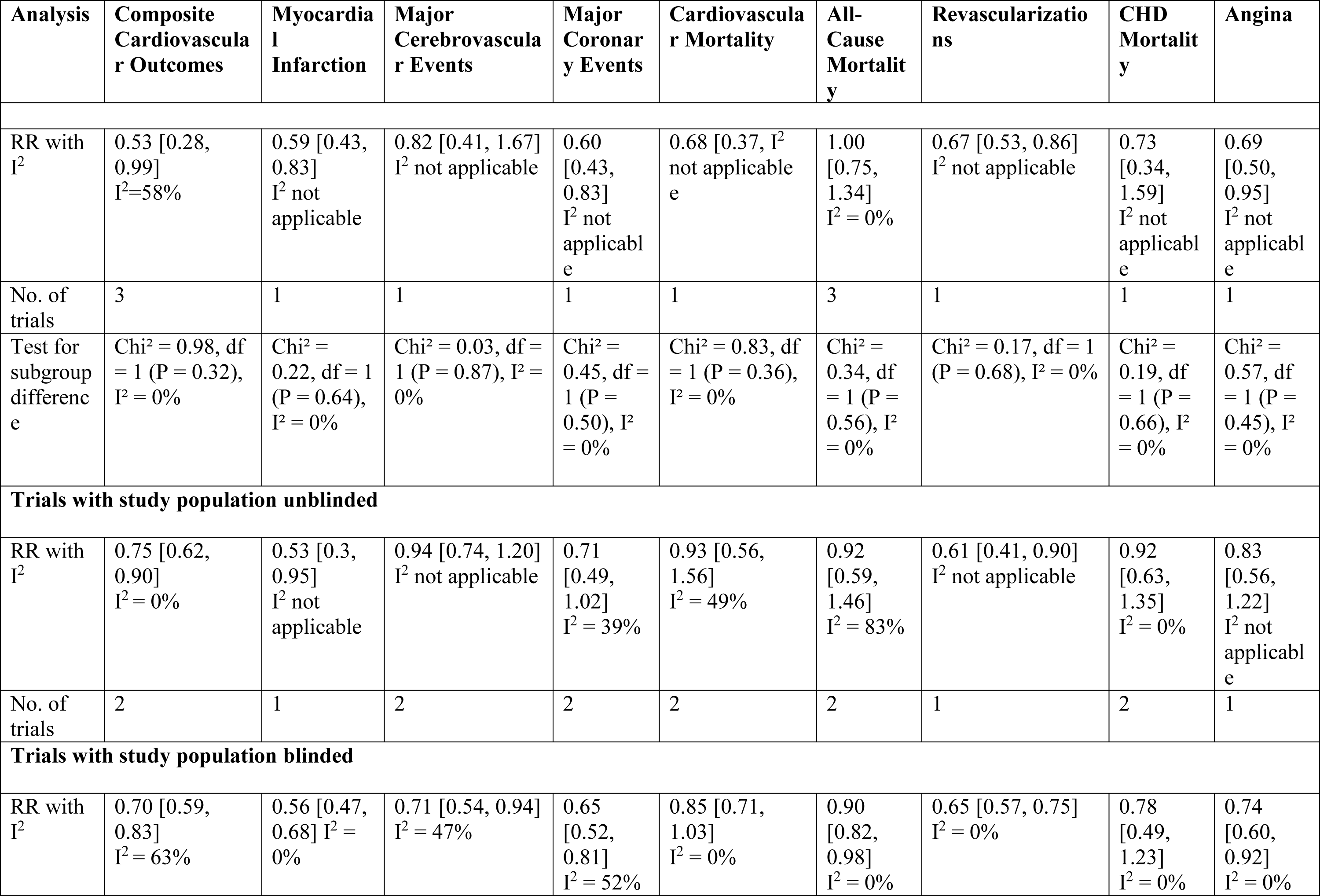
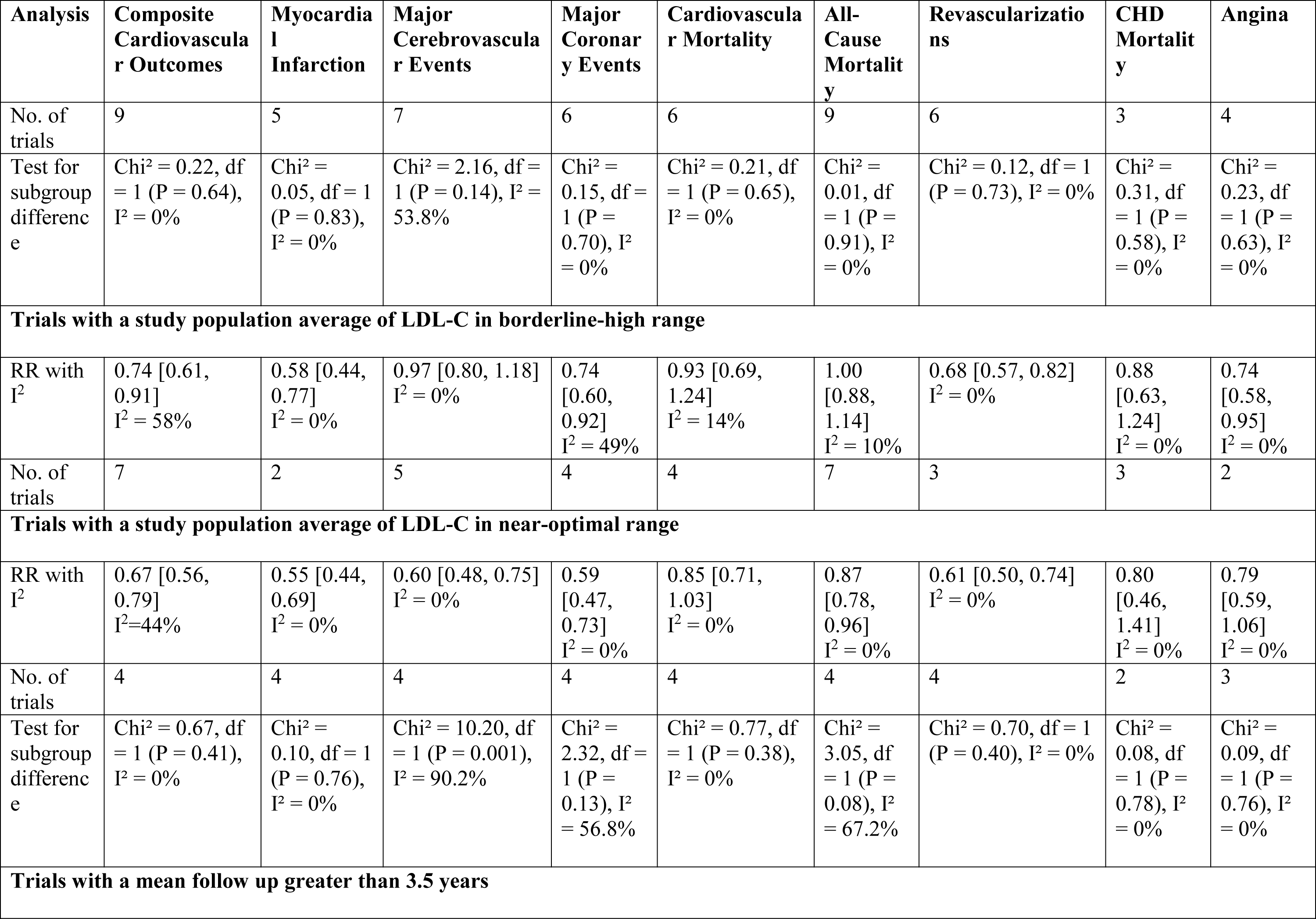
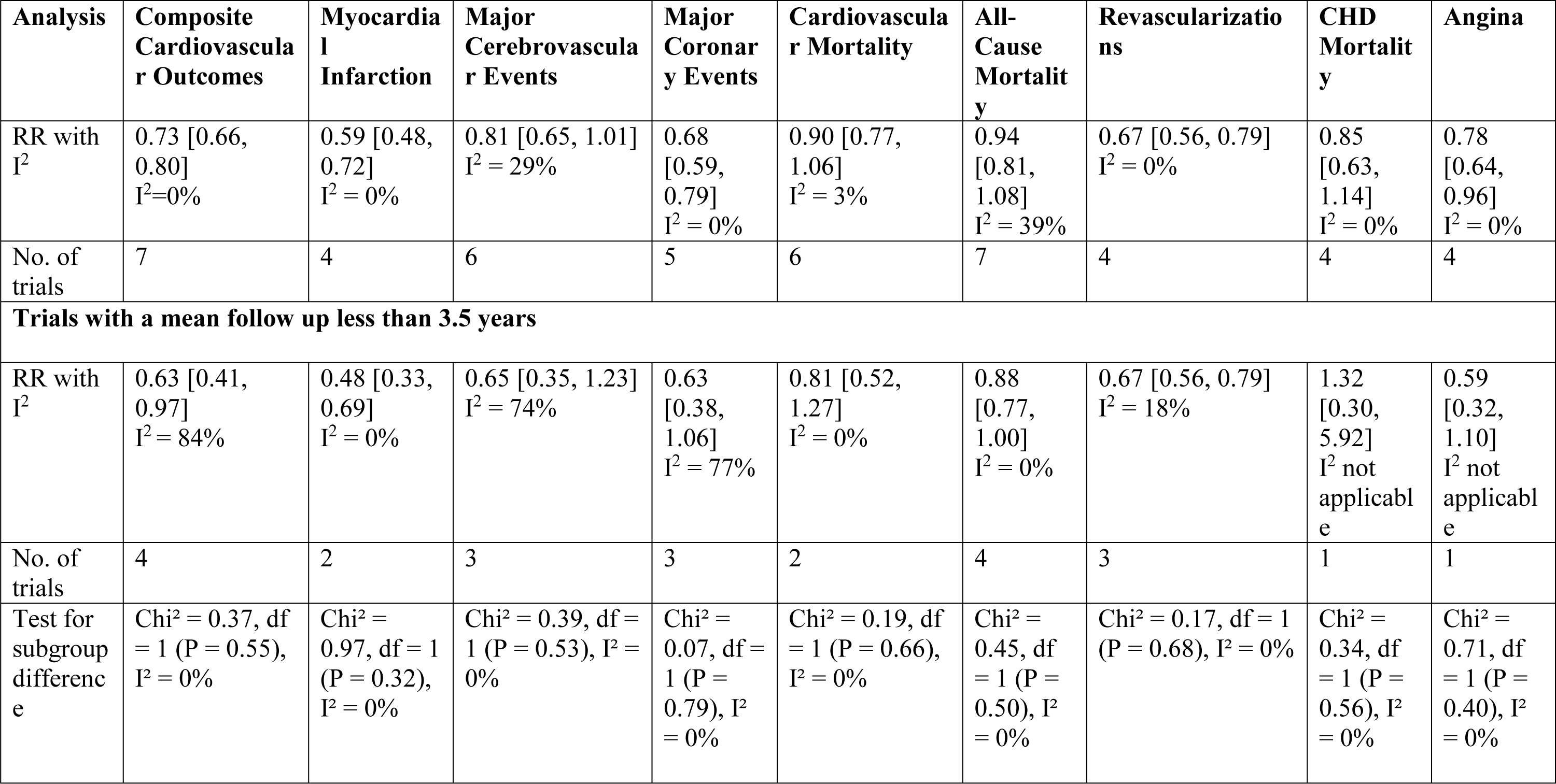
Sensitivity analysis stratified for the trial characteristics

**Supplement Table 4.**
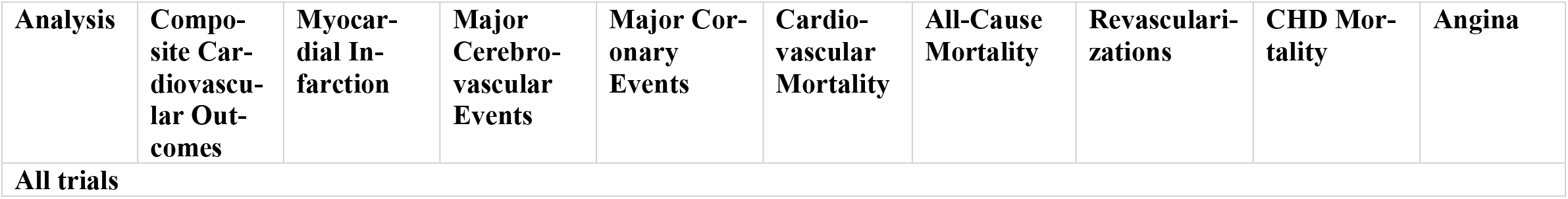
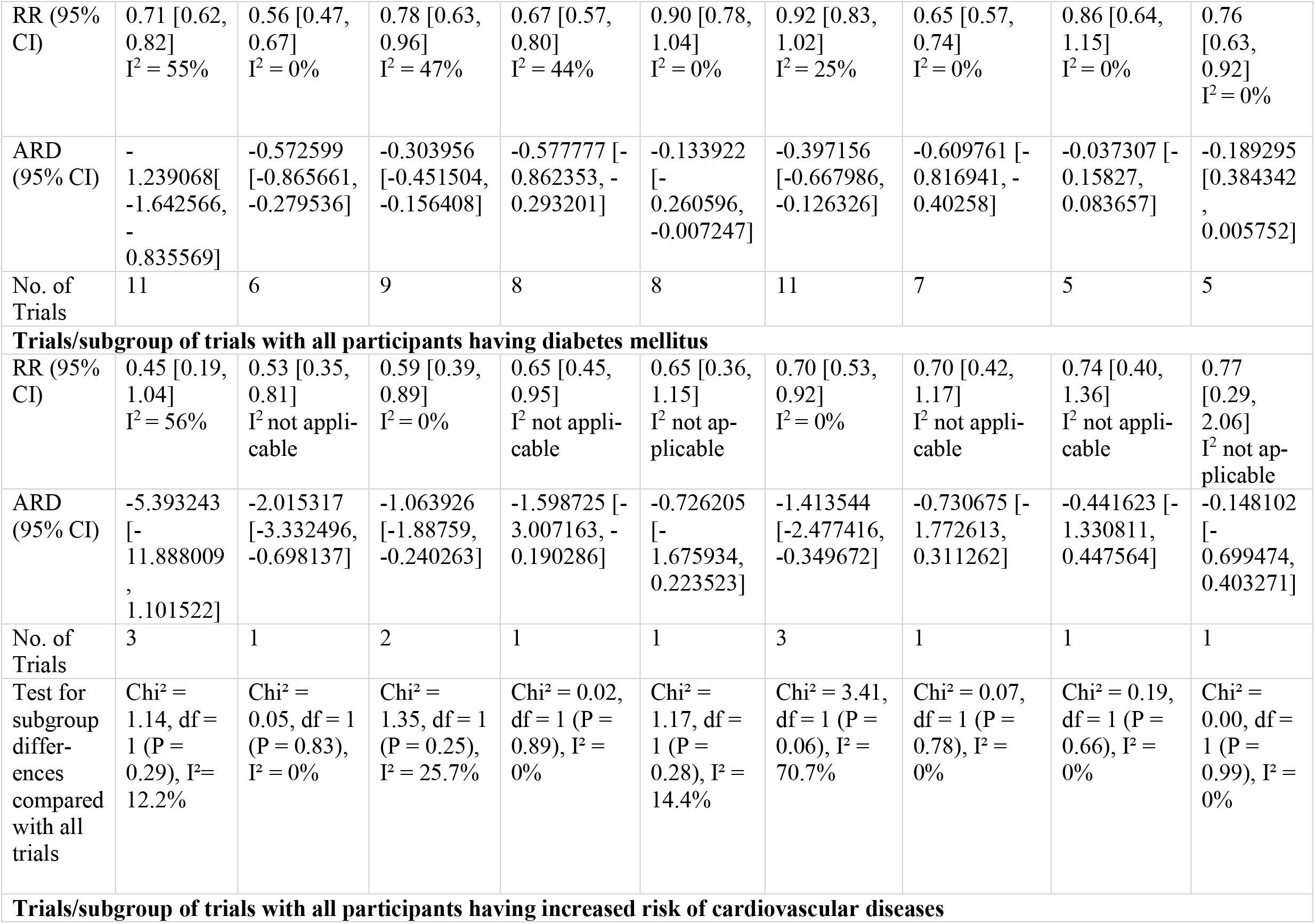
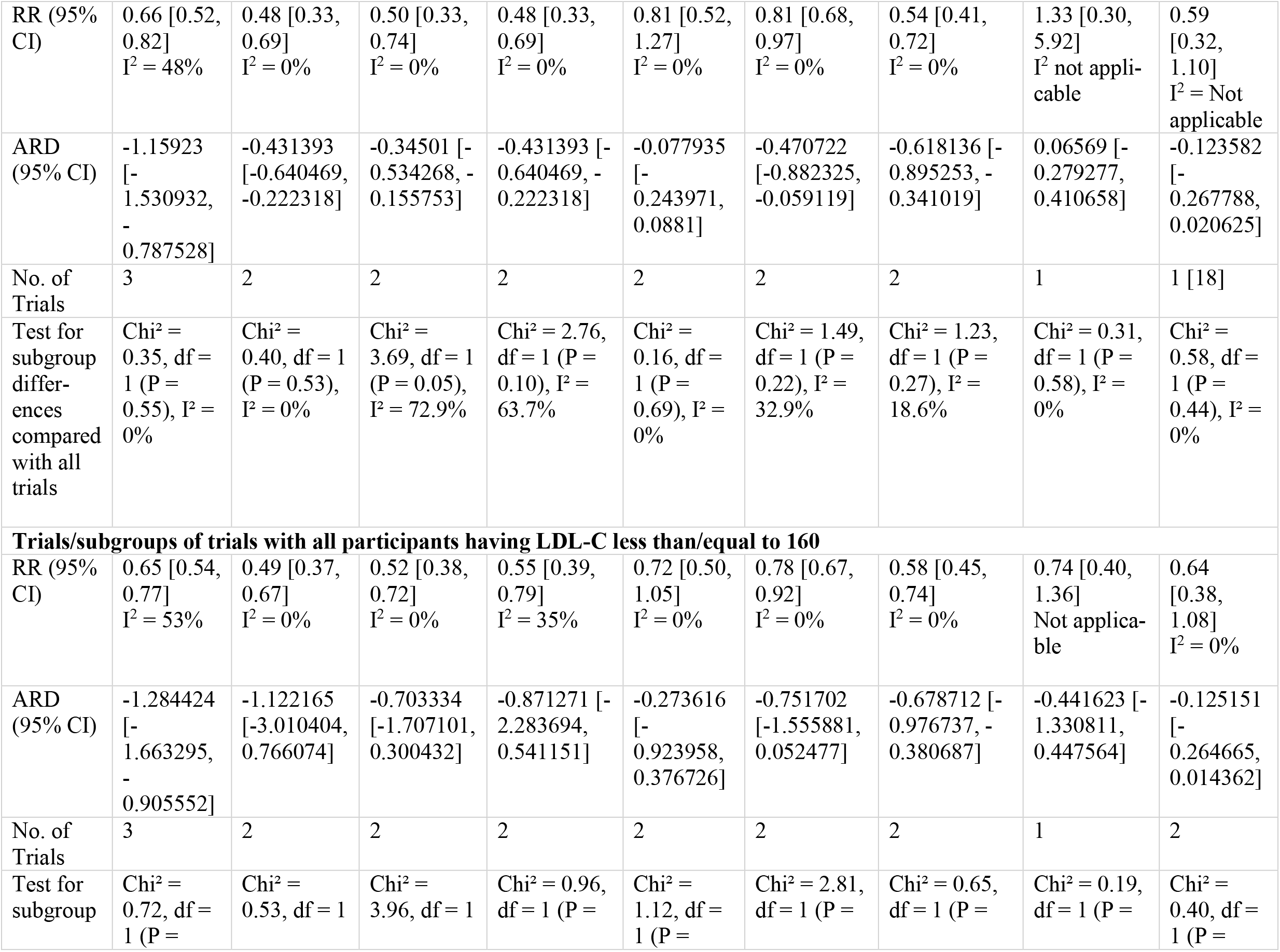
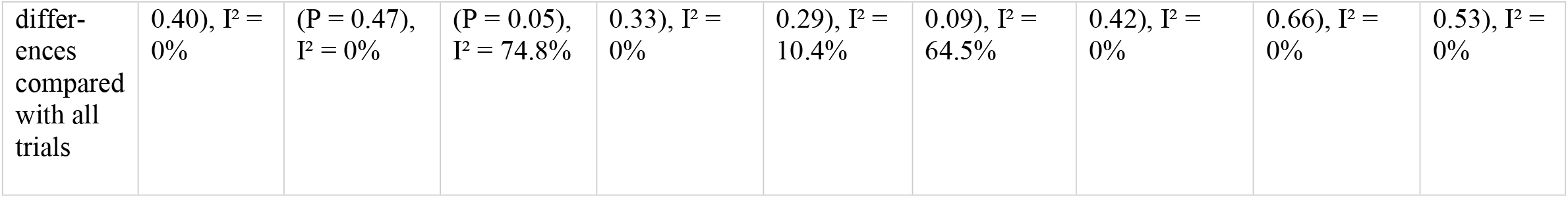
Sensitivity analysis stratified for the type of population

**Supplement Table 5.**
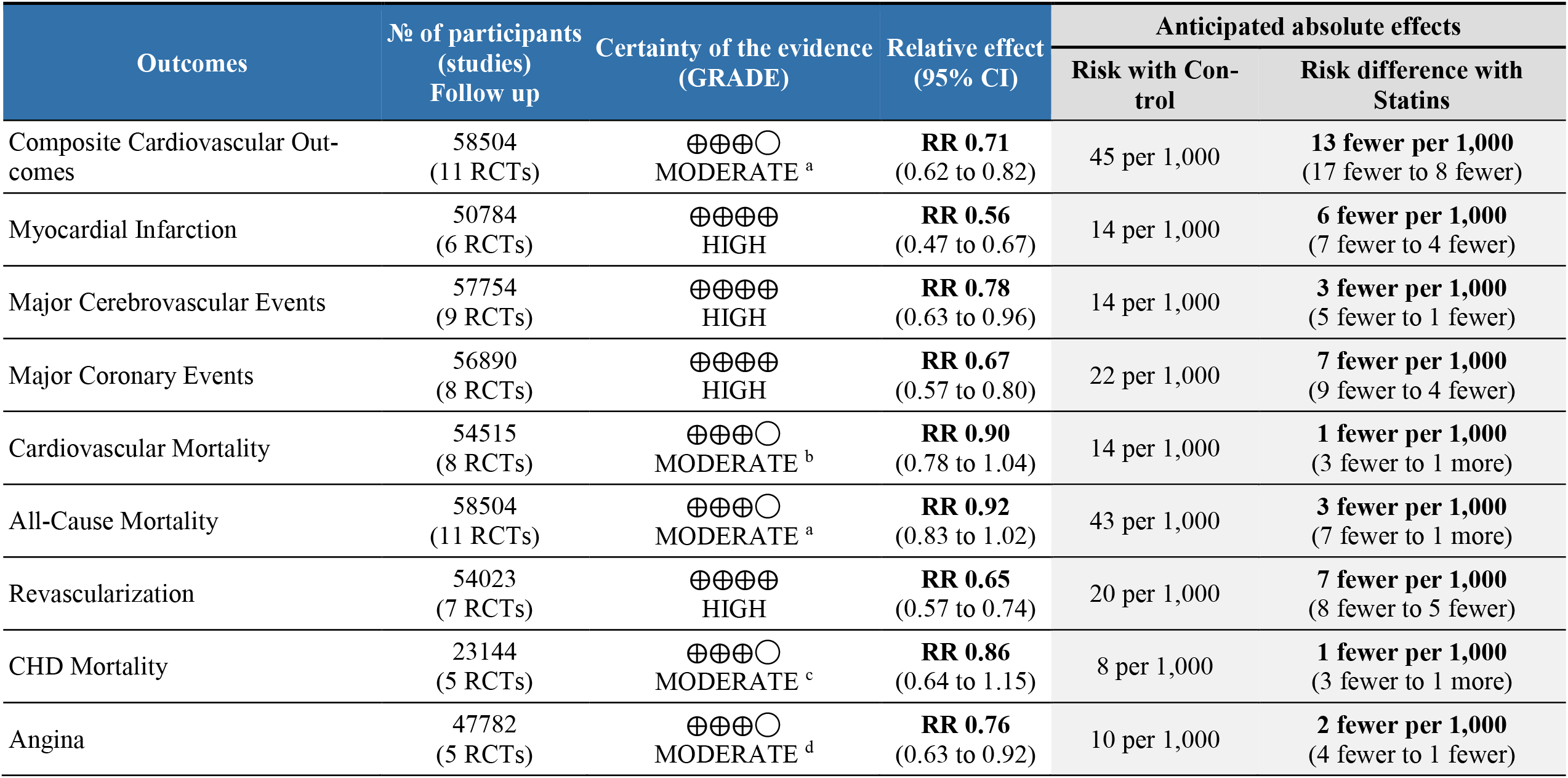
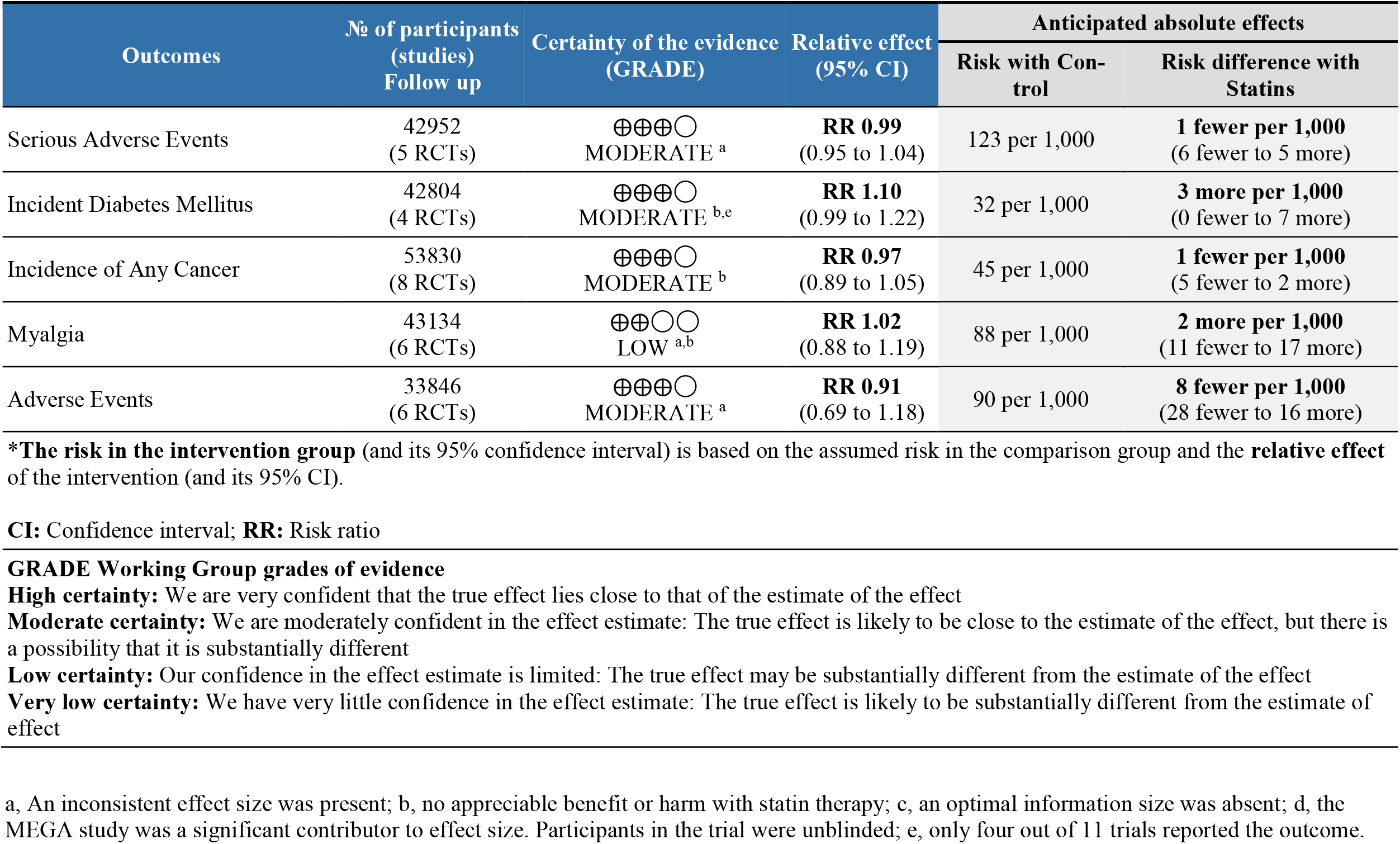
Grade of evidence

| Outcomes | № of participants (studies)<br>Follow up | Certainty of the evidence (GRADE) | Relative effect (95% CI) | Anticipated absolute effects |  |
| --- | --- | --- | --- | --- | --- |
|  |  |  |  | Risk with Control | Risk difference with Statins |
| Composite Cardiovascular Outcomes | 58504<br>(11 RCTs) | ⊕⊕⊕○<br>MODERATE <sup>a</sup> | **RR 0.71**<br>(0.62 to 0.82) | 45 per 1,000 | **13 fewer per 1,000**<br>(17 fewer to 8 fewer) |
| Myocardial Infarction | 50784<br>(6 RCTs) | ⊕⊕⊕⊕<br>HIGH | **RR 0.56**<br>(0.47 to 0.67) | 14 per 1,000 | **6 fewer per 1,000**<br>(7 fewer to 4 fewer) |
| Major Cerebrovascular Events | 57754<br>(9 RCTs) | ⊕⊕⊕⊕<br>HIGH | **RR 0.78**<br>(0.63 to 0.96) | 14 per 1,000 | **3 fewer per 1,000**<br>(5 fewer to 1 fewer) |
| Major Coronary Events | 56890<br>(8 RCTs) | ⊕⊕⊕⊕<br>HIGH | **RR 0.67**<br>(0.57 to 0.80) | 22 per 1,000 | **7 fewer per 1,000**<br>(9 fewer to 4 fewer) |
| Cardiovascular Mortality | 54515<br>(8 RCTs) | ⊕⊕⊕○<br>MODERATE <sup>b</sup> | **RR 0.90**<br>(0.78 to 1.04) | 14 per 1,000 | **1 fewer per 1,000**<br>(3 fewer to 1 more) |
| All-Cause Mortality | 58504<br>(11 RCTs) | ⊕⊕⊕○<br>MODERATE <sup>a</sup> | **RR 0.92**<br>(0.83 to 1.02) | 43 per 1,000 | **3 fewer per 1,000**<br>(7 fewer to 1 more) |
| Revascularization | 54023<br>(7 RCTs) | ⊕⊕⊕⊕<br>HIGH | **RR 0.65**<br>(0.57 to 0.74) | 20 per 1,000 | **7 fewer per 1,000**<br>(8 fewer to 5 fewer) |
| CHD Mortality | 23144<br>(5 RCTs) | ⊕⊕⊕○<br>MODERATE <sup>c</sup> | **RR 0.86**<br>(0.64 to 1.15) | 8 per 1,000 | **1 fewer per 1,000**<br>(3 fewer to 1 more) |
| Angina | 47782<br>(5 RCTs) | ⊕⊕⊕○<br>MODERATE <sup>d</sup> | **RR 0.76**<br>(0.63 to 0.92) | 10 per 1,000 | **2 fewer per 1,000**<br>(4 fewer to 1 fewer) |

| Outcomes | № of participants<br>(studies)<br>Follow up | Certainty of the evidence<br>(GRADE) | Relative effect<br>(95% CI) | Anticipated absolute effects |  |
| --- | --- | --- | --- | --- | --- |
|  |  |  |  | Risk with Con-<br>trol | Risk difference with<br>Statins |
| Serious Adverse Events | 42952<br>(5 RCTs) | ⊕⊕⊕○<br>MODERATE <sup>a</sup> | **RR 0.99**<br>(0.95 to 1.04) | 123 per 1,000 | **1 fewer per 1,000**<br>(6 fewer to 5 more) |
| Incident Diabetes Mellitus | 42804<br>(4 RCTs) | ⊕⊕⊕○<br>MODERATE <sup>b,e</sup> | **RR 1.10**<br>(0.99 to 1.22) | 32 per 1,000 | **3 more per 1,000**<br>(0 fewer to 7 more) |
| Incidence of Any Cancer | 53830<br>(8 RCTs) | ⊕⊕⊕○<br>MODERATE <sup>b</sup> | **RR 0.97**<br>(0.89 to 1.05) | 45 per 1,000 | **1 fewer per 1,000**<br>(5 fewer to 2 more) |
| Myalgia | 43134<br>(6 RCTs) | ⊕⊕○○<br>LOW <sup>a,b</sup> | **RR 1.02**<br>(0.88 to 1.19) | 88 per 1,000 | **2 more per 1,000**<br>(11 fewer to 17 more) |
| Adverse Events | 33846<br>(6 RCTs) | ⊕⊕⊕○<br>MODERATE <sup>a</sup> | **RR 0.91**<br>(0.69 to 1.18) | 90 per 1,000 | **8 fewer per 1,000**<br>(28 fewer to 16 more) |

\***The risk in the intervention group** (and its 95% confidence interval) is based on the assumed risk in the comparison group and the **relative effect** of the intervention (and its 95% CI).
**CI:** Confidence interval; **RR:** Risk ratio

### Supplementary Material 4

Supplement Figure 1-9

**Supplement Figure 1.**
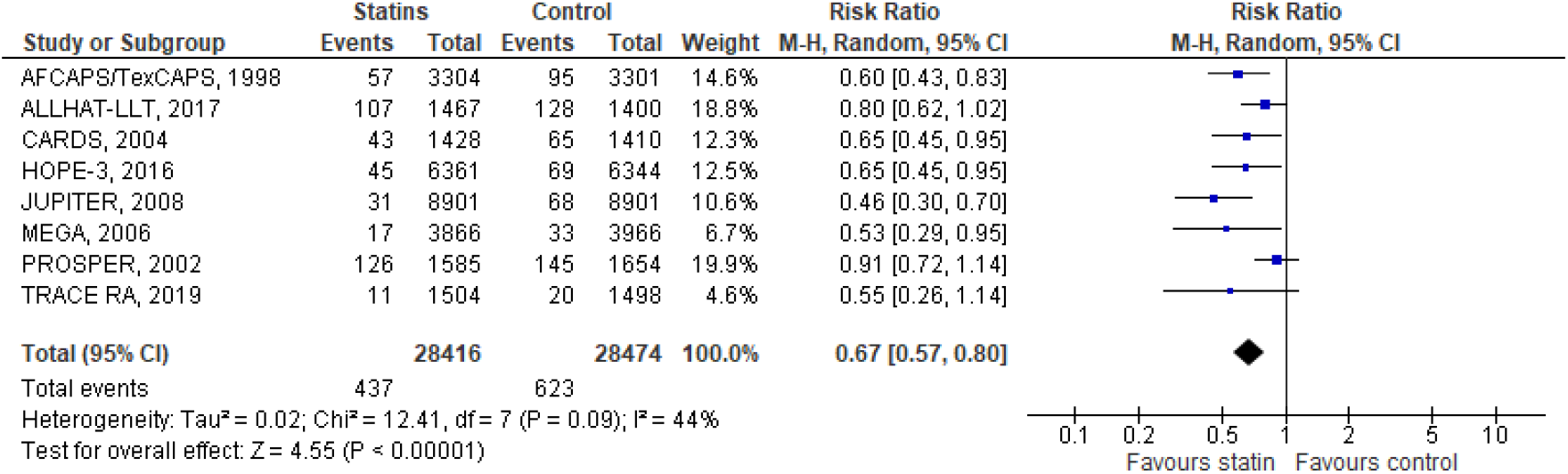
Meta-analysis of major coronary events. AFCAPS/TexCAPS, Air Force/Texas Coronary Atherosclerosis Prevention Study; ALLHAT-LLT, Antihypertensive and Lipid-Lowering Treatment to Prevent Heart Attack; CARDS, Collaborative Atorvastatin Diabetes Study; HOPE–3, Heart Outcomes Prevention Evaluation; JUPITER, Justification for the Use of Statins in Prevention: An Intervention Trial Evaluating Rosuvastatin; MEGA, Management of Elevated Cholesterol in the Primary Prevention Group of Adult Japanese; PROSPER, PROspective Study of Pravastatin in the Elderly at Risk; TRACE RA, Trial of Atorvastatin for the Primary Prevention of Cardiovascular Events in Patients with Rheumatoid Arthritis. In the meta-analysis of major coronary events, we included events reported by trials as follows: AFCAPS/TexCAPS, HOPE-3, JUPITER, and MEGA reported any myocardial infarction (fatal and non-fatal myocardial infarction); ALLHAT-LLT reported fatal coronary heart disease and non-fatal myocardial infarction; CARDS reported fatal myocardial infarction, non-fatal MI, and other acute CHD deaths; PROSPER reported coronary heart disease deaths, and non-fatal myocardial infarction; and TRACE-RA reported non-fatal myocardial infarction.

**Supplement Figure 2.**
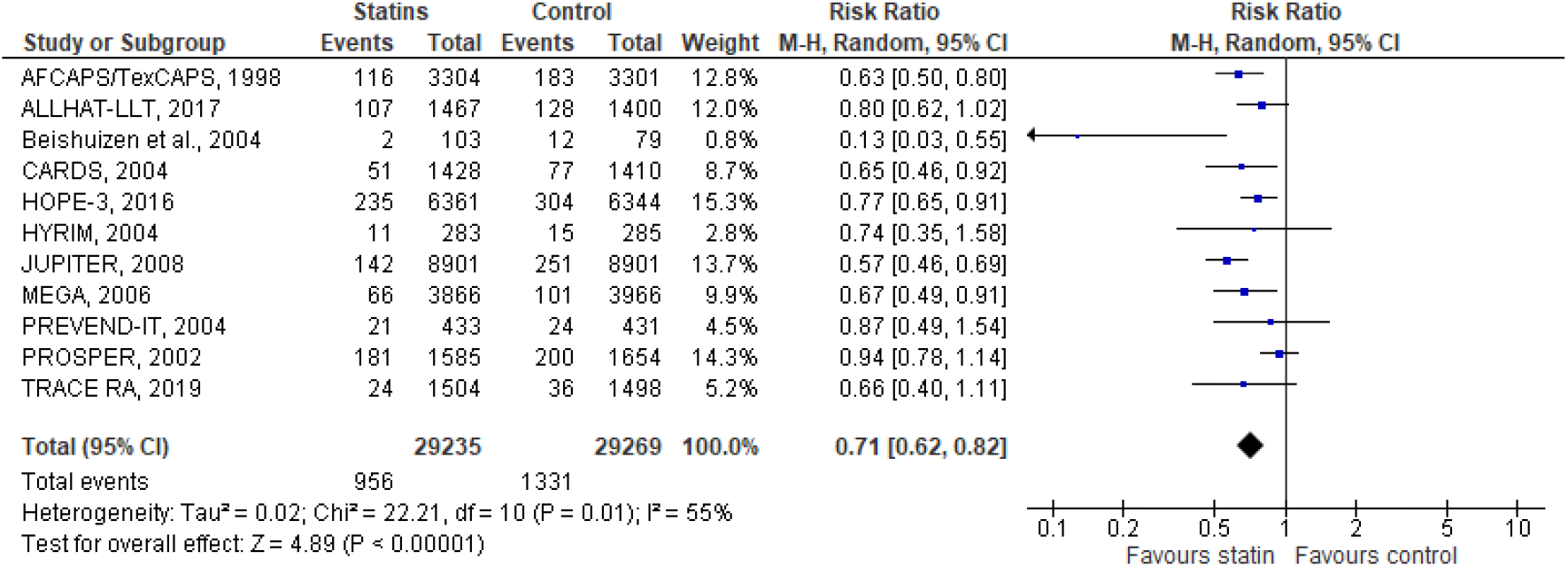
Meta-analysis of composite cardiovascular outcome. AFCAPS/TexCAPS, Air Force/Texas Coronary Atherosclerosis Prevention Study; ALLHAT-LLT, Antihypertensive and Lipid-Lowering Treatment to Prevent Heart Attack; CARDS, Collaborative Atorvastatin Diabetes Study; HOPE–3, Heart Outcomes Prevention Evaluation; HYRIM, Hypertension High Risk Management trial; JUPITER, Justification for the Use of Statins in Prevention: An Intervention Trial Evaluating Rosuvastatin; MEGA, Management of Elevated Cholesterol in the Primary Prevention Group of Adult Japanese; PREVEND IT, the Prevention of REnal and Vascular ENdstage Disease Intervention Trial; PROSPER, PROspective Study of Pravastatin in the Elderly at Risk; TRACE RA, Trial of Atorvastatin for the Primary Prevention of Cardiovascular Events in Patients with Rheumatoid Arthritis. In the meta-analysis of composite cardiovascular outcomes, we included events reported by trials as follows: AFCAPS/TexCAPS reported fatal myocardial infarction, non-fatal myocardial infarction, unstable angina, and sudden cardiac deaths; ALLHAT-LLT reported fatal coronary heart disease and non-fatal myocardial infarction; Beishuizen et al reported unspecified cardiovascular events; CARDS reported myocardial infarction, including silent myocardial infarction, unstable angina, acute coronary heart disease deaths, and resuscitated cardiac arrest; HOPE-3 reported deaths from cardiovascular causes, non-fatal myocardial infarction or non-fatal stroke; HYRIM reported myocardial infarction, sudden death, fatal or non-fatal stroke, transient ischemic attacks, and heart failure; JUPITER reported non-fatal myocardial infarction, non-fatal stroke, cardiovascular mortality, hospitalization for unstable angina, and arterial revascularization procedures; MEGA reported first occurrence of coronary heart disease (fatal or non-fatal myocardial infarction), angina, sudden cardiac deaths, and coronary revascularization procedures; PREVEND IT reported combined incidence of cardiovascular mortality, and hospitalization for cardiovascular morbidity; PROSPER reported coronary heart disease death, non-fatal my-ocardial infarction, and fatal or non-fatal stroke; and TRACE RA reported non-fatal myocardial infarction, non-fatal presumed ischemic stroke, transient ischemic attack, coronary or non-coronary revascularization, cerebrovascular deaths excluding cerebral hemorrhage and non-coronary cardiac death.

**Supplement Figure 3.**
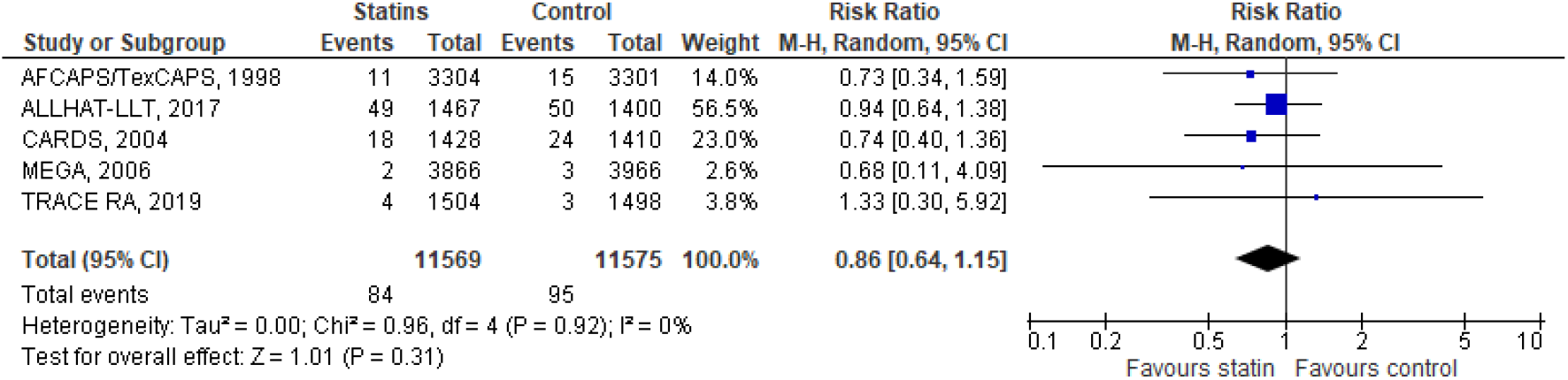
Meta-analysis of coronary heart disease mortality. AFCAPS/TexCAPS, Air Force/Texas Coronary Atherosclerosis Prevention Study; ALLHAT-LLT, Antihypertensive and Lipid-Lowering Treatment to Prevent Heart Attack; CARDS, Collaborative Atorvastatin Diabetes Study; MEGA, Management of Elevated Cholesterol in the Primary Prevention Group of Adult Japanese; TRACE RA, Trial of Atorvastatin for the Primary Prevention of Cardiovascular Events in Patients with Rheumatoid Arthritis. In the meta-analysis of coronary heart disease mortality, we included events reported by trials as follows: AFCAPS/TexCAPS reported fatal coronary heart disease events; ALLHAT-LLT reported coronary heart disease deaths; CARDS reported occurrence of the first event as fatal myocardial infarction or other acute coronary heart disease deaths; MEGA reported fatal myocardial infarction; TRACE-RA reported coronary deaths.

**Supplement Figure 4.**
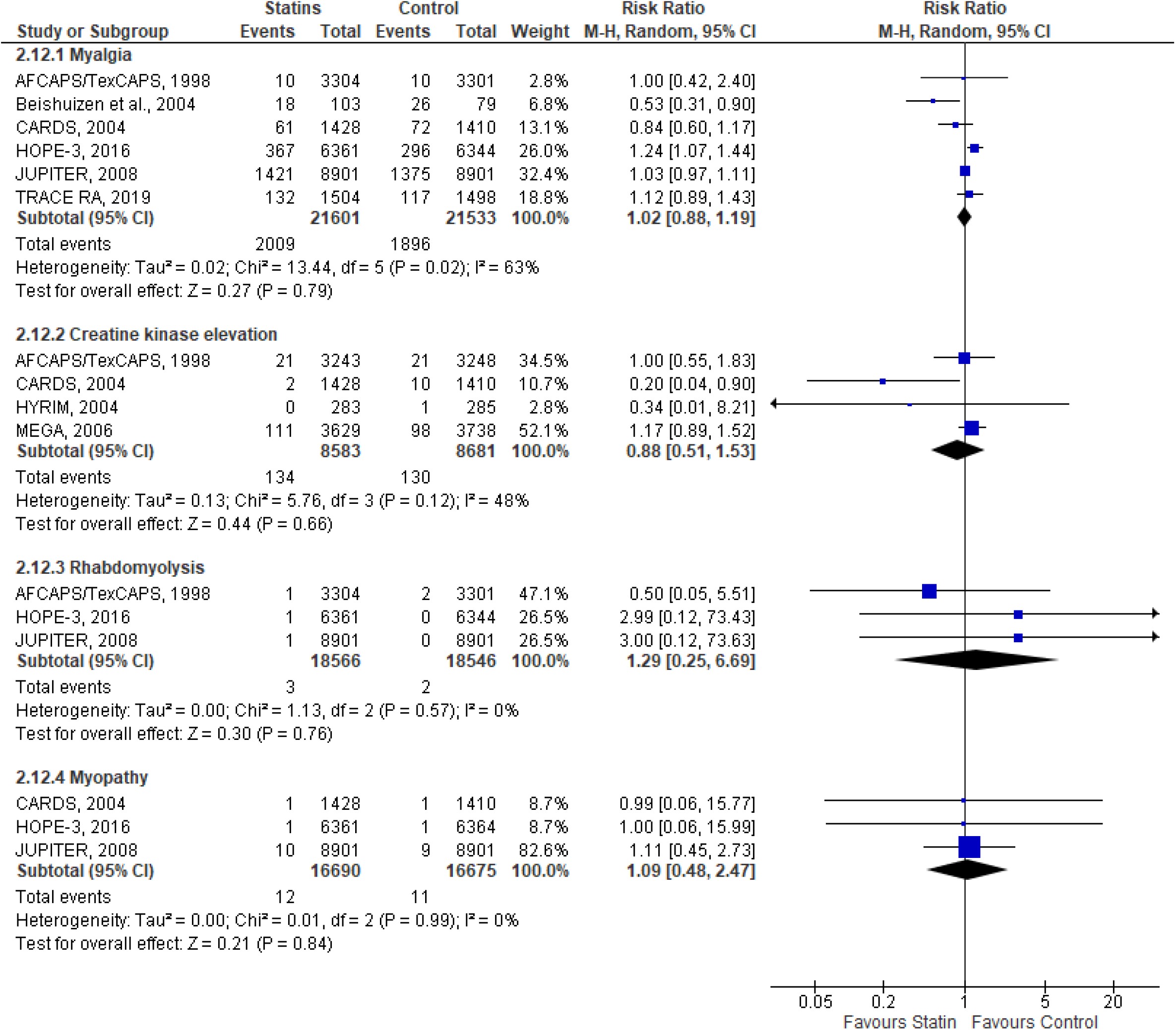
Meta-analysis of muscle-related adverse events. AFCAPS/TexCAPS, Air Force/Texas Coronary Atherosclerosis Prevention Study; CARDS, Collaborative Atorvastatin Diabetes Study; HOPE–3, Heart Outcomes Prevention Evaluation; HYRIM, Hypertension High Risk Management trial; JUPITER, Justification for the Use of Statins in Prevention: An Intervention Trial Evaluating Rosuvastatin; MEGA, Management of Elevated Cholesterol in the Primary Prevention Group of Adult Japanese; TRACE RA, Trial of Atorvastatin for the Primary Prevention of Cardiovascular Events in Patients with Rheumatoid Arthritis.

**Supplement Figure 5.**
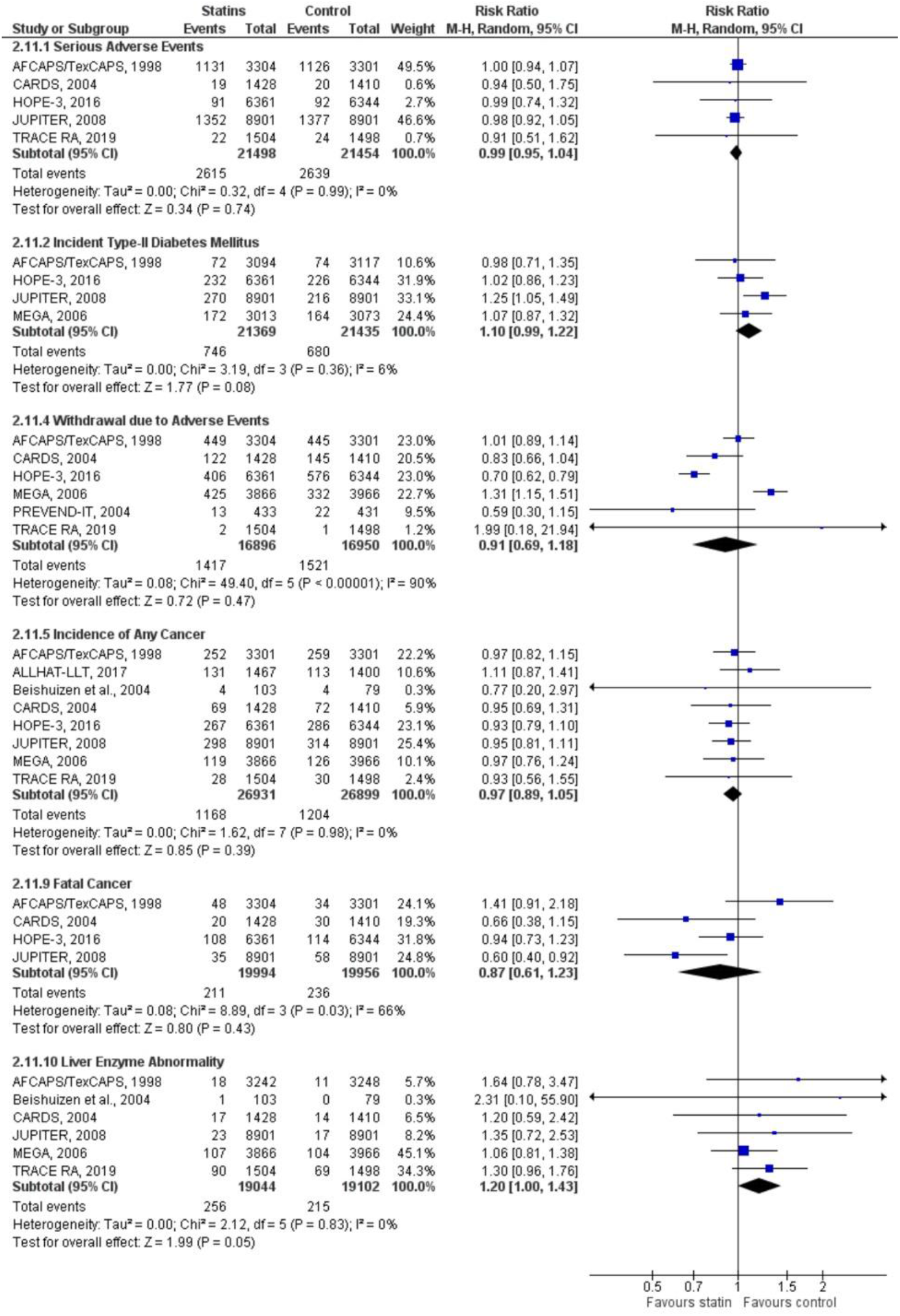
Meta-analysis of incidence of other adverse events. AFCAPS/TexCAPS, Air Force/Texas Coronary Atherosclerosis Prevention Study; ALLHAT-LLT, Antihypertensive and Lipid-Lowering Treatment to Prevent Heart Attack; CARDS, Collaborative Atorvastatin Diabetes Study; HOPE–3, Heart Outcomes Prevention Evaluation; JUPITER, Justification for the Use of Statins in Prevention: An Intervention Trial Evaluating Rosuvastatin; MEGA, Management of Elevated Cholesterol in the Primary Prevention Group of Adult Japanese; TRACE RA, Trial of Atorvastatin for the Primary Prevention of Cardiovascular Events in Patients with Rheumatoid Arthritis.

**Supplement Figure 6.**
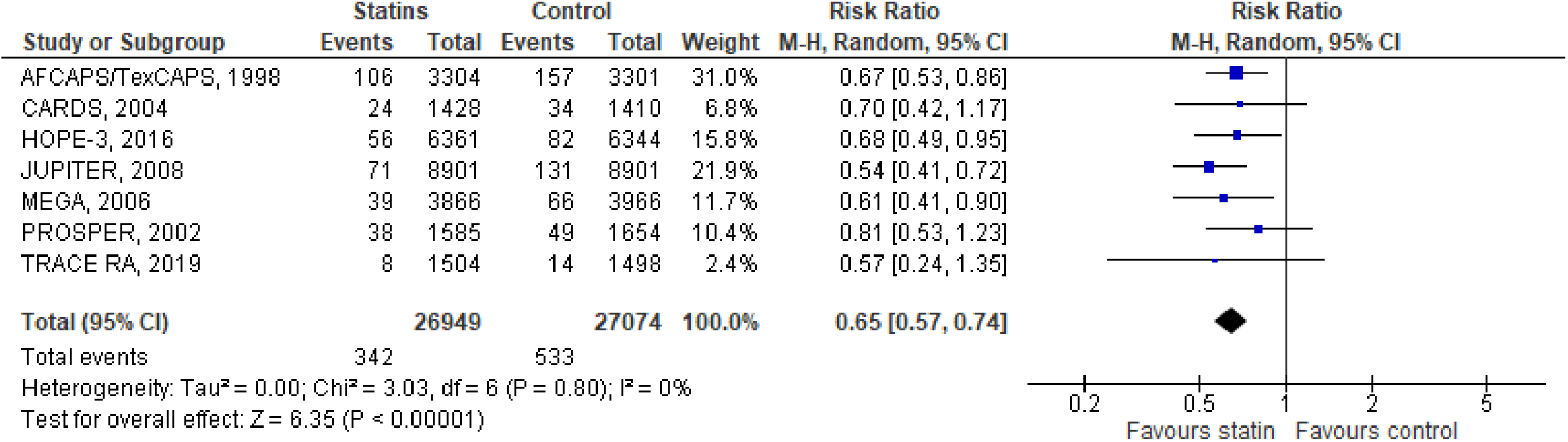
Meta-analysis of revascularizations. AFCAPS/TexCAPS, Air Force/Texas Coronary Atherosclerosis Prevention Study; CARDS, Collaborative Atorvastatin Diabetes Study; HOPE–3, Heart Outcomes Prevention Evaluation; JUPITER, Justification for the Use of Statins in Prevention: An Intervention Trial Evaluating Rosuvastatin; MEGA, Management of Elevated Cholesterol in the Primary Prevention Group of Adult Japanese; PROSPER, PROspective Study of Pravastatin in the Elderly at Risk; TRACE RA, Trial of Atorvastatin for the Primary Prevention of Cardiovascular Events in Patients with Rheumatoid Arthritis. In the meta-analysis of revascularizations, we included events reported by trials as follows: AFCAPS/TexCAPS, HOPE-3, PROSPER reported any revascularizations; CARDS, MEGA, TRACE RA reported coronary revascularizations; JUPITER reported arterial revascularizations.

**Supplement Figure 7.**
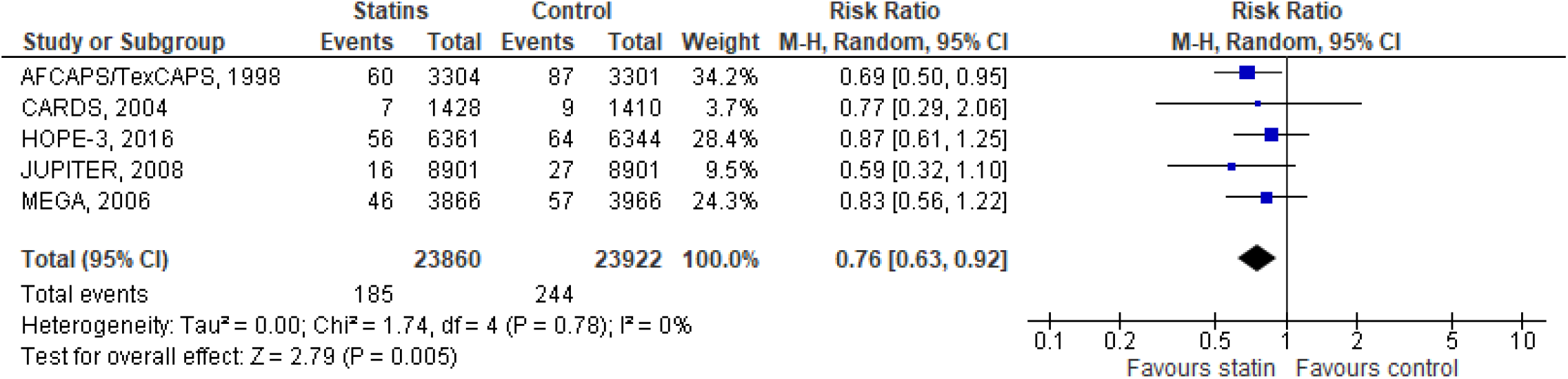
Meta-analysis of angina. AFCAPS/TexCAPS, Air Force/Texas Coronary Atherosclerosis Prevention Study; CARDS, Collaborative Atorvastatin Diabetes Study; HOPE–3, Heart Outcomes Prevention Evaluation; JUPITER, Justification for the Use of Statins in Prevention: An Intervention Trial Evaluating Rosuvastatin; MEGA, Management of Elevated Cholesterol in the Primary Prevention Group of Adult Japanese. In the meta-analysis of angina, we included events reported by trials as follows: AFCAPS/TexCAPS, CARDS, and HOPE-3 reported unstable angina; JUPITER reported hospitalization for unstable angina; and MEGA reported any angina.

**Supplement Figure 8.**
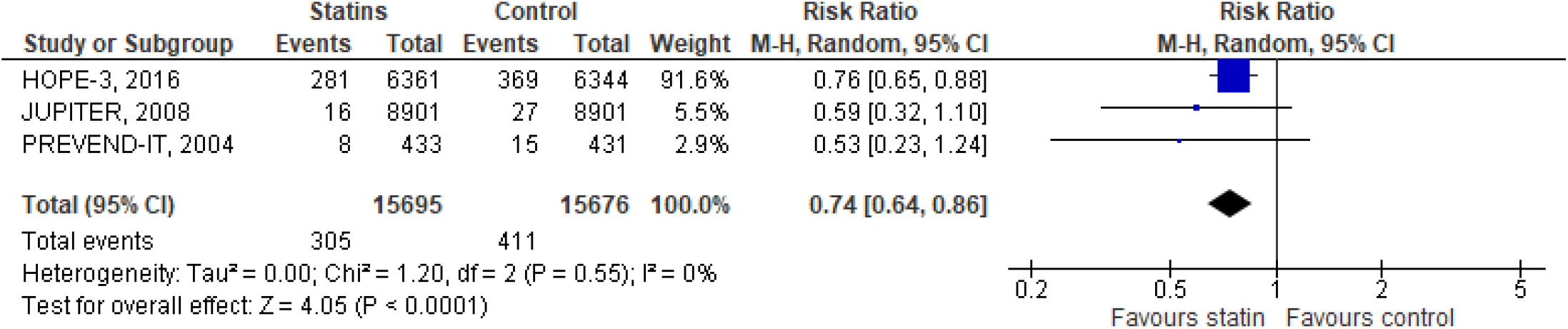
Meta-analysis of hospitalizations for cardiovascular causes. HOPE–3, Heart Outcomes Prevention Evaluation; JUPITER, Justification for the Use of Statins in Prevention: An Intervention Trial Evaluating Rosuvastatin; PREVEND IT, the Prevention of REnal and Vascular ENdstage Disease Intervention Trial. In the meta-analysis of composite cardiovascular outcomes, we included events reported by trials as follows: (1) HOPE-3, 2016 reported hospitalizations for cardiovascular causes; (2) JUPITER, 2008 reported hospitalizations for unstable angina; and (3) PREVEND-IT reported hospitalizations for non-fatal myocardial infarction and myocardial ischemia.

**Supplement Figure 9.**
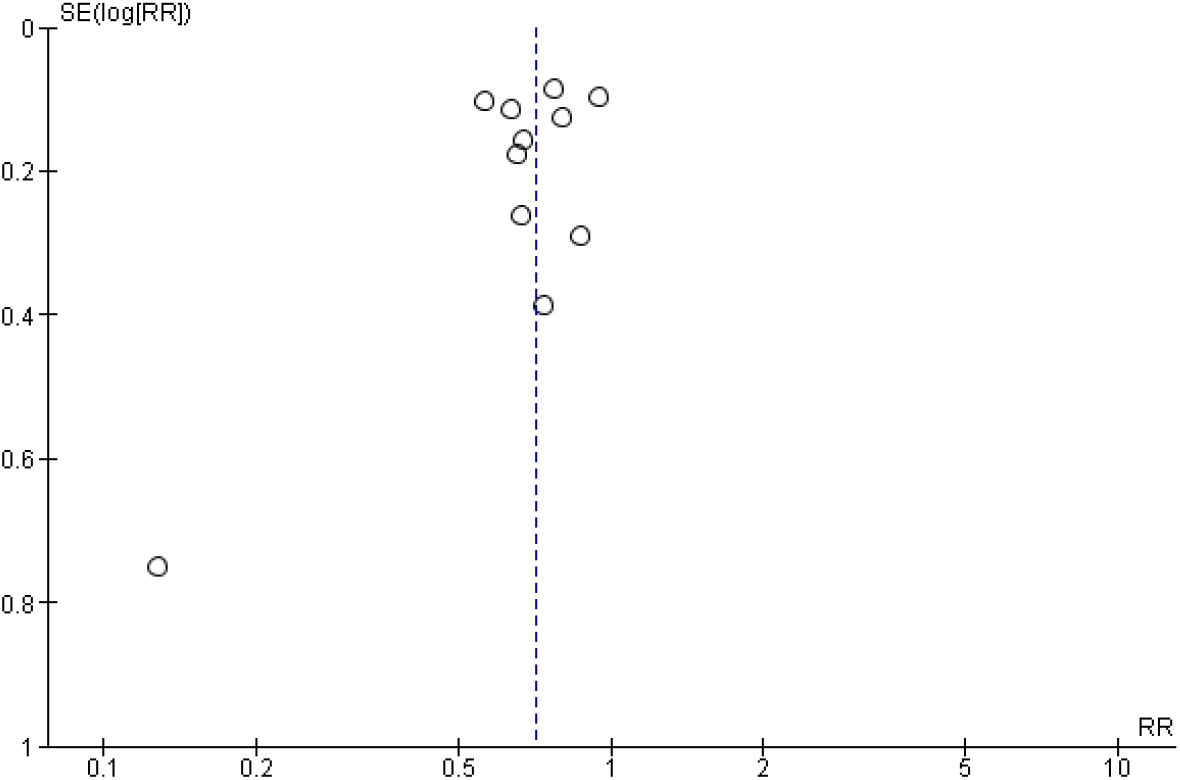
Funnel plot for composite cardiovascular outcomes.

